# Beyond *BRCA* deficiency: Clinical and molecular predictors of survival in patients with *BRCA*-deficient tubo-ovarian high-grade serous carcinoma

**DOI:** 10.1101/2025.09.17.25335919

**Authors:** Tibor A. Zwimpfer, Sian Fereday, Ahwan Pandey, Dinuka Ariyaratne, Madawa W. Jayawardana, Laura Twomey, Céline M. Laumont, Catherine J. Kennedy, Adelyn Bolithon, Nicola S. Meagher, Katy Milne, Phineas Hamilton, Jennifer Alsop, Antonis C. Antoniou, George Au-Yeung, Matthias W. Beckmann, Amy Berrington de Gonzalez, Christiani Bisinotto, Freya Blome, Clara Bodelon, Jessica Boros, Alison H. Brand, Michael E. Carney, Alicia Cazorla-Jiménez, Derek S. Chiu, Elizabeth L. Christie, Anita Chudecka-Głaz, Penny Coulson, Kara L. Cushing-Haugen, Cezary Cybulski, Kathleen M. Darcy, Cath David, Trent Davidson, Arif B. Ekici, Esther Elishaev, Julius Emons, Tobias Engler, Rhonda Farrell, Anna Fischer, Montserrat García-Closas, Aleksandra Gentry-Maharaj, Prafull Ghatage, Rosalind Glasspool, Philipp Harter, Andreas D. Hartkopf, Arndt Hartmann, Sebastian Heikaus, Brenda Y. Hernandez, Anusha Hettiaratchi, Sabine Heublein, David G. Huntsman, Mercedes Jimenez-Linan, Michael E. Jones, Eunyoung Kang, Ewa Kaznowska, Tomasz Kluz, Felix K.F. Kommoss, Gottfried Konecny, Roy F.P.M. Kruitwagen, Jessica Kwon, Diether Lambrechts, Cheng-Han Lee, Jenny Lester, Samuel C.Y. Leung, Yee Leung, Anna Linder, Jolanta Lissowska, Liselore Loverix, Jan Lubiński, Constantina Mateoiu, Iain A. McNeish, Malak Moubarak, Gregg S. Nelson, Nikilyn Nevins, Alexander B. Olawaiye, Siel Olbrecht, Sandra Orsulic, Ana Osorio, Carmel M. Quinn, Ganendra Raj Mohan, Isabelle Ray-Coquard, Cristina Rodríguez-Antona, Patricia Roxburgh, Matthias Ruebner, Stuart G. Salfinger, Spinder Samra, Minouk J. Schoemaker, Hans-Peter Sinn, Gabe S. Sonke, Linda Steele, Colin J.R. Stewart, Aline Talhouk, Adeline Tan, Christopher M. Tarney, Sarah E. Taylor, Koen K. Van de Vijver, Maaike A. van der Aa, Toon Van Gorp, Els Van Nieuwenhuysen, Lilian van-Wagensveld, Andrea E. Wahner-Hendrickson, Christina Walter, Chen Wang, Julia Welz, Nicolas Wentzensen, Lynne R. Wilkens, Stacey J. Winham, Boris Winterhoff, Michael S. Anglesio, Andrew Berchuck, Francisco J. Candido dos Reis, Paul A. Cohen, Thomas P. Conrads, Philip Crowe, Jennifer A. Doherty, Peter A. Fasching, Renée T. Fortner, María J. García, Simon A. Gayther, Marc T. Goodman, Jacek Gronwald, Holly R. Harris, Florian Heitz, Hugo M. Horlings, Beth Y. Karlan, Linda E. Kelemen, G. Larry Maxwell, Usha Menon, Francesmary Modugno, Susan L. Neuhausen, Joellen M. Schildkraut, Annette Staebler, Karin Sundfeldt, Anthony J. Swerdlow, Ignace Vergote, Anna H. Wu, James D. Brenton, Paul D.P. Pharoah, Celeste Leigh Pearce, Malcolm C. Pike, Ellen L. Goode, Susan J. Ramus, Martin Köbel, Brad H. Nelson, Anna DeFazio, Michael L. Friedlander, David D.L. Bowtell, Dale W. Garsed

**Affiliations:** Peter MacCallum Cancer Centre, Melbourne, Victoria, Australia, 3000; Department of Biomedicine, University of Basel, Basel, Switzerland, 4031; Gynecological Cancer Centre, University Hospital Basel, Basel, Switzerland, 4031; The Sir Peter MacCallum Department of Oncology, The University of Melbourne, Parkville, Victoria, Australia, 3010; Deeley Research Centre, BC Cancer. Victoria, BC, Canada; Centre for Cancer Research, The Westmead Institute for Medical Research, Sydney, New South Wales, Australia, 2145; Department of Gynaecological Oncology, Westmead Hospital, Sydney, New South Wales, Australia, 2145; Faculty of Medicine and Health, The University of Sydney, Sydney, New South Wales, Australia, 2006; School of Clinical Medicine, UNSW Medicine and Health, University of NSW Sydney, Sydney, New South Wales, Australia, 2052; Adult Cancer Program, Lowy Cancer Research Centre, University of NSW Sydney, Sydney, New South Wales, Australia, 2052; The Daffodil Centre, The University of Sydney, a joint venture with Cancer Council NSW, Sydney, New South Wales, Australia, 2006; Department of Oncology, University of Cambridge. Cambridge, UK, CB2 0QQ; Centre for Cancer Genetic Epidemiology, Department of Public Health and Primary Care, University of Cambridge, Cambridge, UK, CB1 8RN; Department of Gynecology and Obstetrics, Comprehensive Cancer Center Erlangen-EMN, Friedrich-Alexander University Erlangen-Nuremberg. University Hospital Erlangen, Erlangen, Germany, 91054; Division of Genetics and Epidemiology, The Institute of Cancer Research, London, UK, SW7 3RP; Department of Gynecology and Obstetrics, Ribeirão Preto Medical School, University of São Paulo. Ribeirão Preto, Brazil; Institute of Pathology and Neuropathology, Tuebingen University Hospital, Tuebingen, Germany; Department of Population Science, American Cancer Society, Atlanta, GA, USA, 30303; Department of Obstetrics and Gynecology, John A. Burns School of Medicine, University of Hawaii, Honolulu, HI, USA; Pathology Department, Fundación Jiménez Díaz. Madrid, Spain; British Columbia’s Gynecological Cancer Research Team OVCARE, University of British Columbia, BC Cancer, and Vancouver General Hospital, Vancouver, BC, Canada, V5Z 4E6; Department of Gynecological Surgery and Gynecological Oncology of Adults and Adolescents, Pomeranian Medical University, Szczecin, Poland, 70-111; Program in Epidemiology, Division of Public Health Sciences, Fred Hutchinson Cancer Center, Seattle, WA, USA, 98109; Department of Comparative Biomedical Sciences, College of Veterinary Medicine, Mississippi State University, Mississippi State, MS, USA, 39762; Gynecologic Cancer Center of Excellence, Department of Gynecologic Surgery and Obstetrics, Uniformed Services University of the Health Sciences, Walter Reed National Military Medical Center, Bethesda, MD, USA; Henry M Jackson Foundation for the Advancement of Military Medicine, Inc. Bethesda, MD, USA; Gynaecological Cancer Centre, Royal Hospital for Women, Randwick, New South Wales, Australia; NSW Health Pathology, Prince of Wales Hospital. Sydney, New South Wales, Australia; School of Medicine, Western Sydney University, Penrith, New South Wales, Australia, 2751; Institute of Human Genetics. Comprehensive Cancer Center Erlangen-EMN, University Hospital Erlangen, Friedrich-Alexander University Erlangen-Nuremberg FAU, Erlangen, Germany, 91054; Department of Pathology, University of Pittsburgh School of Medicine, Pittsburgh, PA, USA; Department of Women’s Health, Tuebingen University Hospital, Tuebingen, Germany, 72076; Prince of Wales Private Hospital, Randwick, New South Wales, Australia; MRC Clinical Trials Unit, Institute of Clinical Trials and Methodology, University College London, London, UK, WC1V 6LJ; Department of Women’s Cancer, Elizabeth Garrett Anderson Institute for Women’s Health, University College London, London, UK; Department of Oncology, Division of Gynecologic Oncology, Cumming School of Medicine, University of Calgary. Calgary, AB, Canada; Beatson West of Scotland Cancer Centre and School of Cancer Sciences, University of Glasgow, Glasgow, UK, G12 0YN; Department of Gynecology and Gynecologic Oncology, Evangelische Kliniken Essen-Mitte, Essen, Germany, 45136; Department of Gynecology and Obstetrics, University Hospital of Ulm, Ulm, Germany, 89075; Institute of Pathology, Comprehensive Cancer Center Erlangen-EMN, Friedrich-Alexander University Erlangen-Nuremberg, University Hospital Erlangen, Erlangen, Germany, 91054; Center for Pathology, Evangelische Kliniken Essen-Mitte, Essen, Germany; University of Hawaii Cancer Center, Honolulu, HI, USA; The Health Precincts Biobank, UNSW Biospecimen Services, Mark Wainwright Analytical Centre, UNSW, Sydney, New South Wales, Australia; Department of Obstetrics and Gynecology, University Hospital Heidelberg, Heidelberg, Germany; Department of Obstetrics and Gynecology, University of British Columbia, Vancouver, BC, Canada, V5Z 1L3; Department of Molecular Oncology, BC Cancer Research Centre, Vancouver, BC, Canada, V5Z 4E6; Department of Histopathology, Addenbrooke’s Hospital, Cambridge, UK; Department of Surgery, Seoul National University Bundang Hospital, Seongnam, Korea, 13260; Department of Pathology, Institute of Medical Sciences, Medical College of Rzeszow University, Rzeszow, Poland; Department of Gynecology, Gynecology Oncology and Obstetrics, Institute of Medical Sciences, Medical College of Rzeszów University, Rzeszów, Poland; Institute of Pathology, Heidelberg University Hospital, Heidelberg, Germany; David Geffen School of Medicine, Department of Obstetrics and Gynecology, University of California at Los Angeles, Los Angeles, CA, USA, 90095; Department of Obstetrics and Gynecology, Maastricht University Medical Centre, Maastricht, the Netherlands; GROW Ð School for Oncology and Reproduction, Maastricht University Medical Center, Maastricht, the Netherlands, 6229 HX; Laboratory for Translational Genetics, Department of Human Genetics, KU Leuven, Leuven, Belgium, 3000; VIB Center for Cancer Biology, VIB Leuven, Belgium, 3001; Department of Pathology and Laboratory Medicine, University of Alberta, Edmonton, Alberta, Canada; Department of Gynaecological Oncology, King Edward Memorial Hospital, Subiaco, Western Australia, Australia; Division of Obstetrics and Gynaecology, Medical School, University of Western Australia, Crawley, Western Australia, Australia; Department of Obstetrics and Gynecology, Institute of Clinical Science, Sahlgrenska Center for Cancer Research, University of Gothenburg, Gothenburg, Sweden; Department of Cancer Epidemiology and Prevention, M Sklodowska-Curie National Research Oncology Institute, Warsaw, Poland, 02-034; Division of Gynecologic Oncology, Department of Gynecology and Obstetrics. Leuven Cancer Institute. Leuven, Belgium, 3000; Department of Genetics and Pathology, Pomeranian Medical University, Szczecin, Poland, 71-252; Department of Pathology, University of Gothenburg, Gothenburg, Sweden; Division of Cancer and Ovarian Cancer Action Research Centre, Department Surgery & Cancer, Imperial College London, London, UK, W12 0NN; School of Cancer Sciences, University of Glasgow, Glasgow, UK, G61 1QH; Division of Gynecologic Oncology, Department of Obstetrics, Gynecology and Reproductive Sciences, University of Pittsburgh School of Medicine, Pittsburgh, PA, USA, 15213; Genetics Service, Fundación Jiménez Díaz University Hospital and Health Research Institute, Universidad Autónoma de Madrid IIS-FJD, UAM, Madrid, Spain, 28040; Centre for Biomedical Network Research on Rare Diseases CIBERER, Instituto de Salud Carlos III, Madrid, Spain, 28029; Department of Gynaecological Oncology, St John of God Subiaco Hospital, Subiaco, Western Australia, Australia; Centre Leon Berard and University Claude Bernard Lyon 1, Lyon, France, 69373; Pharmacogenomics and Tumor Biomarkers Group, Institute for Biomedical Research Sols-Morreale CSIC-UAM, Madrid, Spain, 28029; Tissue Pathology and Diagnostic Oncology, Westmead Hospital, Sydney, New South Wales, Australia; Department of Medical Oncology, Netherlands Cancer Institute, Amsterdam, the Netherlands, 1066 CX; Department of Population Sciences, Beckman Research Institute of City of Hope, Duarte, CA, USA, 91010; Gynaepath WA, Clinipath Sonic Healthcare, Osbourne Park, Australia; Department of Pathology, Ghent University Hospital, Cancer Research Institute Ghent CRIG, Ghent, Belgium; Department of Pathology, Antwerp University Hospital, Antwerp, Belgium; Department of Research, Netherlands Comprehensive Cancer Organization IKNL, Utrecht, the Netherlands; Division of Gynaecological Oncology, Leuven Cancer Institute, University Hospital Leuven and KU Leuven, Leuven, Belgium; Department of Oncology, Mayo Clinic, Rochester, MN, USA, 55905; Department of Quantitative Health Sciences, Division of Computational Biology, Mayo Clinic, Rochester, MN, USA, 55905; Division of Cancer Epidemiology and Genetics, National Cancer Institute, Bethesda, MD, USA, 20892; Department of Obstetrics, Gynecology and Women’s Health, University of Minnesota, Minneapolis, MN, USA, 55455; Department of Obstetrics and Gynecology, Division of Gynecologic Oncology, Duke University Medical Center, Durham, NC, USA, 27710; Women’s Health Integrated Research Center, Women’s Service Line, Inova Health System, Falls Church, VA, USA; Huntsman Cancer Institute, Department of Population Health Sciences, University of Utah, Salt Lake City, UT, USA, 84112; Division of Cancer Epidemiology, German Cancer Research Center DKFZ, Heidelberg, Germany, 69120; Department of Research, Cancer Registry of Norway, Norwegian Institute of Public Health, Oslo, Norway; Genomic Biomarkers and Precision Oncology Group, Sols-Morreale Biomedical Research Institute IIBM, Consejo Superior de Investigaciones Cientficas & Universidad Autónoma de Madrid CSIC-UAM, Madrid, Spain, 28029; Medicine, University of Texas Health, San Antonio, San Antonio, USA, 78229; Cancer Prevention and Control Program, Cedars-Sinai Cancer, Cedars-Sinai Medical Center, Los Angeles, CA, USA, 90048; Department of Epidemiology, University of Washington School of Public Health, Seattle, WA, USA, 98195; Department of Gynecology and Gynecological Oncology, HSK, Dr Horst-Schmidt Klinik, Wiesbaden, Wiesbaden, Germany; Department of Pathology, The Netherlands Cancer Institute - Antoni van Leeuwenhoek hospital, Amsterdam, the Netherlands, 1066 CX; Communicable Disease Epidemiology Section, South Carolina Department of Public Health, Columbia, SC, USA; Department of Epidemiology, University of Pittsburgh School of Public Health, Pittsburgh, PA, USA, 15213; Women’s Cancer Research Center, Magee-Womens Research Institute and Hillman Cancer Center, Pittsburgh, PA, USA, 15213; Department of Epidemiology, Rollins School of Public Health, Emory University, Atlanta, GA, USA, 30322; Division of Breast Cancer Research, The Institute of Cancer Research, London, UK, SW7 3RP; Department of Population Health and Public Health Sciences, Keck School of Medicine, University of Southern California Norris Comprehensive Cancer Center, Los Angeles, CA, USA, 90033; Cancer Research UK Cambridge Institute, University of Cambridge, Cambridge, UK, CB2 0RE; Department of Computational Biomedicine, Cedars-Sinai Medical Center, West Hollywood, CA, USA, 90069; Department of Epidemiology, University of Michigan School of Public Health, Ann Arbor, MI, USA, 48109; Department of Epidemiology and Biostatistics, Memorial Sloan-Kettering Cancer Center, New York, NY, USA, 10065; Department of Quantitative Health Sciences, Division of Epidemiology, Mayo Clinic, Rochester, MN, USA, 55905; Department of Pathology and Laboratory Medicine, University of Calgary, Foothills Medical Center, Calgary, AB, Canada, T2N 2T9; Department of Biochemistry and Microbiology, University of Victoria, Victoria, BC, Canada; Department of Medical Genetics, University of British Columbia, Victoria, BC, Canada; Nelune Comprehensive Cancer Centre, Prince of Wales Hospital, Sydney, New South Wales, Australia; Prince of Wales Clinical School, UNSW Medicine and Health, University of NSW Sydney, Sydney, New South Wales, Australia, 2052

## Abstract

*BRCA*-associated homologous recombination deficiency (HRD) is present in ∼50% of high-grade serous carcinomas (HGSC) and predicts sensitivity to platinum-based therapy. However, there is little understanding of why some patients with *BRCA*-deficient tumors experience unexpectedly poor outcomes. We profiled 154 tumors, enriched for patients with *BRCA*-deficient tumors that experienced short overall survival (≤3 years, n=42), using whole-genome, transcriptome, and methylation analyses. All but one *BRCA*-deficient tumor exceeded an accepted HRD genomic scarring threshold. However, patients with *BRCA1*-deficient HGSC with a more elevated HRD score survived significantly longer. Patients with *BRCA2*-deficient HGSC and loss of *NF1* survived twice as long as those without *NF1* loss, whereas *PIK3CA* or *RAD21* amplification defined *BRCA2*-deficient HGSC with exceptionally short survival. *BRCA1*-deficient tumors in short survivors had evidence of immunosuppressive c-kit signaling and EMT. In a large HGSC cohort (n=1,389) including 282 individuals with pathogenic germline *BRCA* variants (g*BRCA*pv), the location of the mutation within functional domains stratified clinical outcomes. Notably, residual disease after primary surgery had limited prognostic effect in g*BRCA*pv-carriers compared to non-carriers. Our findings indicate that tumor HR proficiency in the context of therapy response and survival is not a binary property, and highlight genomic and immune modifiers of outcomes in *BRCA*-deficient HGSC.

## INTRODUCTION

The identification of clinical and molecular determinants of survival in patients with cancer has the dual benefits of finding biomarkers that may guide patient management or provide novel therapeutic opportunities. Until relatively recently, the identification of prognostic biomarkers in ovarian cancer has been confounded by a lack of appreciation of the distinctly different molecular characteristics of the various histologic subtypes that make up epithelial ovarian cancer^1^. Evaluating histologically homogenous sets of ovarian tumors has been critical in deciphering the prognostic importance of proteins such as p53^2,3^ and WT1^4^, and identifying genetic risk loci^5–12^.

High-grade serous carcinoma (HGSC) is the most common histotype, accounting for approximately 70% of ovarian cancer deaths in Western countries^13–16^. Homologous recombination-mediated DNA repair deficiency (HRD) is frequent in HGSC and is most often associated with mutations in *BRCA1* and *BRCA2*^17–19^. Approximately fifty percent of HGSC are regarded to have HRD, a feature that can be inferred through specific patterns of genomic scarring in tumor cells^13,20–25^. HRD leads to genomic instability and tumorigenesis, providing a vulnerability in tumor cells with increased sensitivity to double-strand DNA breaks that can be exploited therapeutically^26–28^. As a result, platinum-based chemotherapy and poly (ADP-ribose) polymerase inhibitor (PARPi) maintenance therapy are generally more effective in patients with HRD tumors^28–33^.

While HRD status is informative, accurate prediction of treatment response and survival in HGSC cannot be simply determined by the presence or absence of mutations in genes associated with HR DNA repair. The initial survival advantage for carriers of pathogenic germline *BRCA1* variants (g*BRCA1*pv) diminishes over time, with fewer g*BRCA1*pv-carriers surviving 10 years after diagnosis than either g*BRCA2*pv-carriers or non-carriers^33–35^. Factors associated with survival outcome in HGSC include residual disease following cytoreductive surgery^16,36–38^, the molecular subtype of the tumor^39^, age at diagnosis^40^, and the extent of T- and B-cell infiltration into tumors^41,42^. In germline pathogenic variant carriers, the location of mutations within *BRCA1* or *BRCA2* or the retention of the wildtype allele in the tumor can result in a hypomorphic phenotype associated with resistance to platinum-based therapy^43–47^. Furthermore, revertant mutations restoring *BRCA1* and *BRCA2* function contribute to acquired resistance to platinum-based therapy and PARPis, impacting treatment response and patient outcomes^48–50^.

Comparing patients who represent the extremes of survival outcomes may provide increased sensitivity to identify prognostic biomarkers that are relevant to a wider patient population^51^. Using this approach, we have recently shown that plasma cell infiltration and other molecular changes, including co-loss of *BRCA* and the tumor suppressor *RB1*, are associated with especially long-term survival in HGSC^22,52,53^. The current study evaluates *BRCA*-deficient HGSC by first focusing on g*BRCA*pv-carriers and then expanding to include somatic mutations and promoter methylation in *BRCA1/2,* and other key HR genes, as well as evaluating tumor HRD status. We focus on patients with either poor or favorable survival outcomes, harnessing the value of analyzing patients with exceptional survival outcomes while comparing cohorts that are as similar as possible in other respects.

## RESULTS

### Association of residual disease with prognosis is attenuated in g*BRCA*pv-carriers

Pathogenic germline *BRCA* variants (g*BRCA*pv) were identified in 20% of patients in the Australian Ovarian Cancer Study (AOCS) cohort (*n*=282/1389) (Table 1, Supplementary Tables S1 and S2). In applying a survival model, there was evidence that the proportional hazards assumption did not hold (*P*<0.001), thus an Accelerated Failure Time (AFT) model^54^ was used with results reported as Time Ratios (TR; see Methods), where TR > 1 indicates longer time to progression or death, and a TR < 1 indicates shorter survival or time to progression. Patients with g*BRCA*pvs exhibited improved overall survival (OS; TR: 1.53, 95% CI: 1.33-1.76, *P*<0.001) and progression-free survival (PFS; TR: 1.34 95% CI: 1.28-1.53, *P*<0.001) compared with non-carriers (Supplementary Tables S3 and S4).

**Table 1:** Baseline characteristics of the clinicopathological features from patients with HGSC of the Australian Ovarian Cancer Study (AOCS) cohort. *G2=Grade 2, G3=Grade 3, OS=Overall survival, gBRCApv=pathogenic germline BRCA variant*

| Characteristics | n=1,389<br>n (%) |
| --- | --- |
| **Age at diagnosis (years)** |  |
| Median | 61 |
| Range | 24-87 |
| Unknown | 7 (0.5) |
| **Germline *BRCA* status** |  |
| Wildtype | 1,107 (79.7) |
| *gBRCA1*pv | 175 (12.6) |
| *gBRCA2*pv | 107 (7.7) |
| **Grade** |  |
| G3 | 1,100 (79.2) |
| G2 | 237 (17.1) |
| Unknown | 52 (3.7) |
| **FIGO stage** |  |
| III-IV | 1,193 (85.9) |
| I-II | 134 (9.6) |
| Unknown | 62 (4.5) |
| **Primary site** |  |
| Ovary | 1,008 (72.6) |
| Peritoneum | 215 (15.5) |
| Fallopian tube | 140 (10.1) |
| Unknown | 26 (1.9) |
| **Surgery** |  |
| Primary cytoreductive surgery | 991 (71.3) |
| Interval cytoreductive surgery | 299 (21.5) |
| Other | 70 (5) |
| Unknown | 29 (2.1) |
| **Residual disease status** |  |
| Residual disease | 829 (59.7) |
| No residual disease | 467 (33.6) |
| Unknown | 93 (6.7) |
| **Neoadjuvant chemotherapy** |  |
| No | 1,060 (76.3) |
| Yes | 322 (23.2) |
| Unknown | 7 (0.5) |
| **PARP inhibitor 1<sup>st</sup> line** |  |
| No | 1,350 (97.2) |
| Yes | 39 (2.8) |
| **Progression-free survival (months)** |  |
| Median | 15 |
| Range | 0-285 |
| Unknown | 11 (0.8) |
| **Overall survival (months)** |  |
| Median | 38 |
| Range | 1-290 |
| Unknown | 11 (0.8) |
| **Status** |  |
| Deceased | 984 (70.8) |
| Alive | 393 (28.3) |
| Unknown | 12 (0.9) |

We considered whether clinical characteristics differed by germline *BRCA* status and found a statistically significant interaction with residual disease status (*P*-interaction=0.011; Supplementary Table S3). Using this interaction term, we found that the negative effect of residual disease after cytoreductive surgery on OS was less pronounced in g*BRCA*pv-carriers (TR: 0.87, 95% CI: 0.72-1.06, *P*=0.162) than in non-carriers (TR: 0.51, 95% CI: 0.44-0.59, *P*<0.001; Fig. 1a, Table 2). The importance of residual disease for survival in non-carriers was confirmed in the independent OTTA cohort (*n*=1004, g*BRCA*pv-carriers=221, 22%; Fig. 1b, Extended Data Figs. 1 and 2a).

**Fig. 1.**
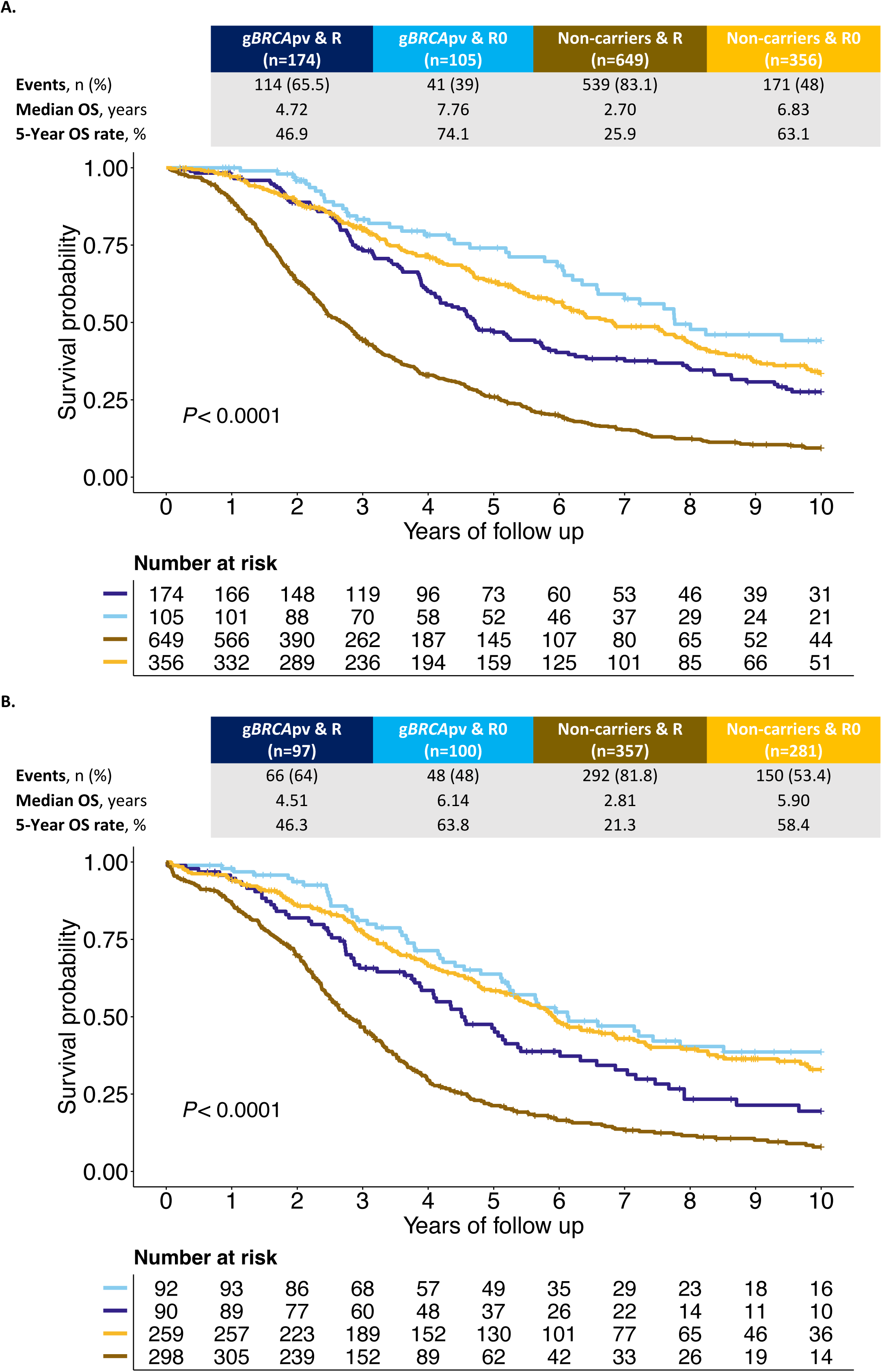
*BRCA* status and residual disease as predictors of overall survival in HGSC. Kaplan-Meier survival curve for the interaction term *BRCA* and Residual status from patients of **a**, the Australian Ovarian Cancer Study (AOCS) cohort and **b**, the Ovarian Tumor Tissue Consortium (OTTA) cohort. *P* values calculated by log-rank test. *R=Residual disease, R0=No residual disease, gBRCApv=pathogenic germline BRCA variant, n=Number of patients, OS=Overall survival*

**Table 2:**
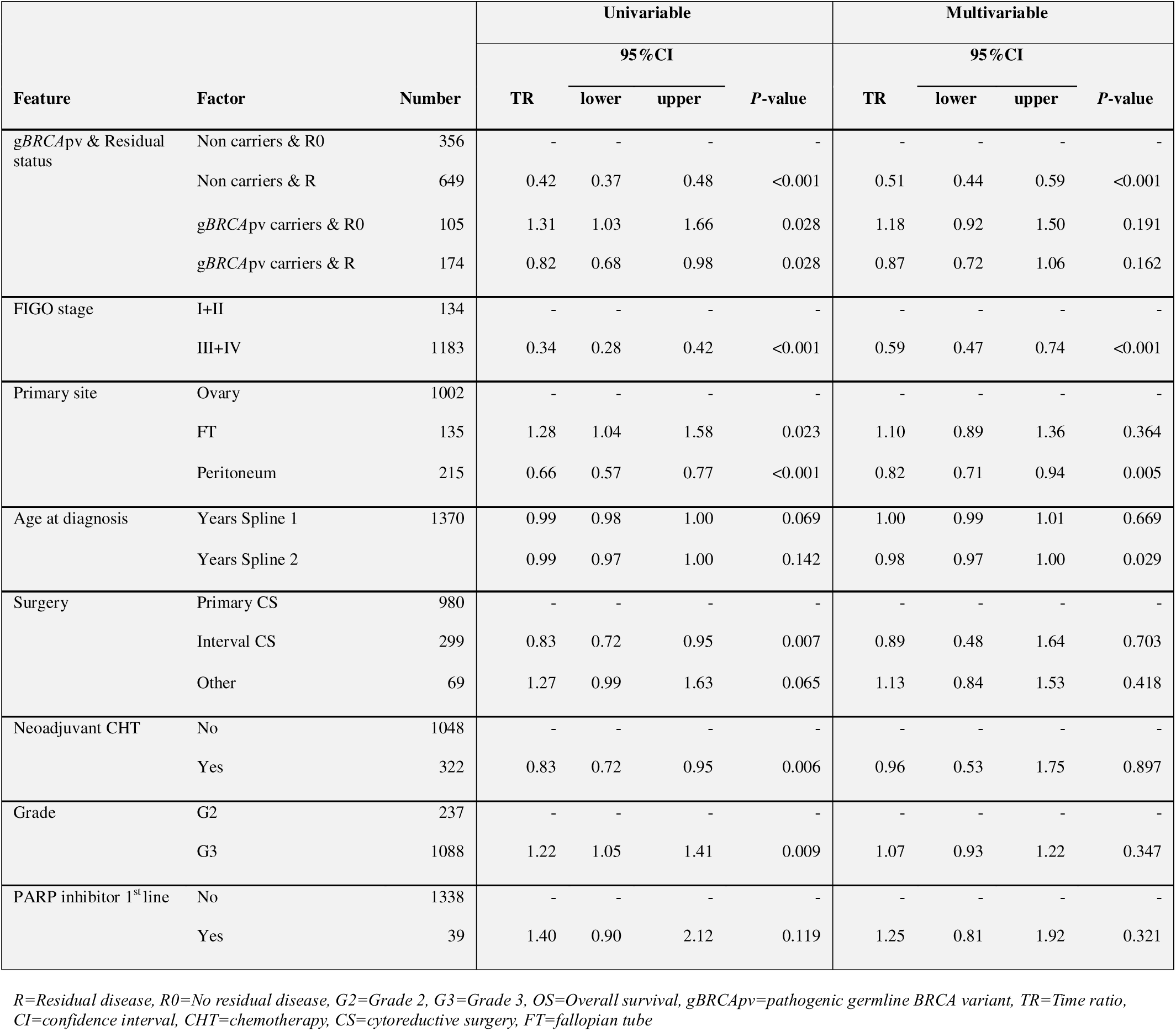
Multivariable Accelerated Failure Time (AFT) model of *BRCA* and residual disease status and clinicopathological predictive features on overall survival in patients from the AOCS cohort. The model was fitted using a log-logistic distribution. Results are expressed as Time Ratios (TR) with corresponding 95% confidence intervals (CI) and *P*-values derived from Wald tests. A TR > 1 indicates a longer survival time, whereas a TR < 1 indicates a shorter survival time. Age at diagnosis was modeled using restricted cubic splines with 3 knots and is presented as two spline terms.

We examined the relationship of residual disease and *BRCA* status to known immune and molecular features associated with survival, including tumor-infiltrating lymphocytes (TIL)^42,55^, *RB1* loss^22,52,56^, and transcriptional molecular subtypes^39^. Non-carriers with residual disease had an inverse association with high CD8+ TIL density (*P*=0.016), with 38.3% of tumors classified as having low or no TIL (Extended Data Fig. 2b, Supplementary Table S5). This group also showed an inverse association with the C4/differentiated (C4.DIF) molecular subtype (*P*=0.010; Extended Data Fig. 2b). We observed an association between the C1/mesenchymal (C1.MES) molecular subtype and residual disease as previously reported^57^, but this was only statistically significant among non-carriers (*P*=0.005). *RB1* loss was associated with g*BRCA*pv-carriers without residual disease (*P*<0.001; Extended Data Fig. 2b).

Although no statistically significant interaction between neoadjuvant chemotherapy (NACT) and *BRCA* status was observed (*P*-interaction=0.12; Supplementary Table S3), there was evidence of heterogeneity of effect in these subgroups. Among participants who did not receive NACT, g*BRCA*pv-carriers showed a survival benefit compared to non-carriers (TR: 1.60, 95% CI: 1.37-1.87, *P*<0.001; Supplementary Table S6, Extended Data Fig. 3). In contrast, the overall survival benefit in g*BRCA*pv-carriers versus non-carriers was not statistically significant in the NACT group (TR: 1.39 and 1.17, 95% CI: 0.75-2.60 and 0.62-2.21, *P*=0.298 and *P*=0.634 respectively, compared to non-carriers who did not receive NACT).

### g*BRCA*pv location and type are associated with survival and therapy response

Mutations located in various functional domains of *BRCA1* and *BRCA2* have been associated with differences in survival and responses to PARPi in ovarian cancer^43,44^. The mutation type and location of g*BRCA*pvs was ascertained for 240 of the patients in the AOCS cohort from their clinical records and/or previous sequencing analyses^22,56,58,59^ (Extended Data Figs. 4a,b and Supplementary Table S2). Following adjustment for FIGO stage, residual disease status, primary site, age, and first-line treatment, patients with g*BRCA1*pvs in exon 10 had a statistically significant improved OS and PFS (TR: 1.54 and 1.49, 95% CI: 1.19-2.00 and 1.16-1.91, *P*<0.001 and *P*=0.002, respectively), but the association was attenuated for those with variants outside exon 10 (TR: 1.21 and 1.18, 95% CI: 0.97-1.51 and 0.96-1.46, *P*=0.09 and *P*=0.12, respectively) compared to non-carriers (Table 3). More specifically, pathogenic variants in the DNA binding domain (DBD) of *BRCA1*, located in exon 10, were associated with an OS and PFS benefit compared to non-carriers (TR: 1.60 and 1.58, 95% CI: 1.14-2.25 and 1.15-2.18, *P*=0.005 and *P*=0.006, respectively; Table 3). In contrast, the OS and PFS benefit was not statistically significant for patients with pathogenic variants in the Really Interesting New Gene (RING) (TR: 1.28 and 1.15, 95% CI: 0.87-1.90 and 0.82-1.61, *P*=0.216 and *P*=0.419, respectively) and C-terminal domains of *BRCA1* (BRCT) (TR: 1.35 and 1.43, 95% CI: 0.83-2.20 and 0.90-2.26, *P*=0.222 and *P*=0.126, respectively), located outside of exon 10.

**Table 3:** Multivariable Accelerated Failure Time (AFT) model analysis of germline *BRCA* pathogenic variant (g*BRCA*pv) location and progression-free survival and overall survival in patients from the AOCS cohort. Models were adjusted for FIGO stage, residual disease status, primary tumor site, type of surgery, age at diagnosis (modeled with restricted cubic splines, 3 knots), use of neoadjuvant chemotherapy, tumor grade, and PARP inhibitor use in first-line treatment. AFT models were fitted using a log-logistic distribution. Results are presented as Time Ratios (TR) with 95% confidence intervals (CI) and *P*-values derived from Wald tests. A TR > 1 indicates an association with longer time to progression or death, while a TR < 1 reflects shorter survival. The reference group for all comparisons is non-carriers of g*BRCA*pv.

| Feature | Factor | Number | Progression-free survival |  |  |  | Overall survival |  |  |  |
| --- | --- | --- | --- | --- | --- | --- | --- | --- | --- | --- |
|  |  |  | TR | 95%CI |  | *P*-value | TR | 95%CI |  | *P*-value |
|  |  |  |  | lower | upper |  |  | lower | upper |  |
| g*BRCA*pv exon | Non carriers | 1096 | - | - | - | - | - | - | - | - |
|  | g*BRCA1*pv Exon 10 | 68 | 1.49 | 1.16 | 1.91 | 0.002 | 1.54 | 1.19 | 2.00 | <0.001 |
|  | g*BRCA1*pv outside Exon 10 | 81 | 1.18 | 0.96 | 1.46 | 0.12 | 1.21 | 0.97 | 1.51 | 0.093 |
|  | g*BRCA2*pv Exon 11 | 52 | 1.52 | 1.16 | 2.00 | 0.002 | 1.67 | 1.26 | 2.23 | <0.001 |
|  | g*BRCA2*pv outside Exon 11 | 38 | 1.66 | 1.15 | 2.42 | 0.007 | 1.90 | 1.31 | 2.76 | <0.001 |
| g*BRCA*pv domain | Non carriers | 1096 | - | - | - | - | - | - | - | - |
|  | g*BRCA1*pv BRCT | 17 | 1.43 | 0.90 | 2.26 | 0.126 | 1.35 | 0.83 | 2.20 | 0.222 |
|  | g*BRCA1*pv DBD | 40 | 1.58 | 1.15 | 2.18 | 0.005 | 1.60 | 1.14 | 2.25 | 0.006 |
|  | g*BRCA1*pv outside domain | 54 | 1.41 | 1.09 | 1.81 | 0.007 | 1.38 | 1.06 | 1.79 | 0.017 |
|  | g*BRCA1*pv RING | 27 | 1.15 | 0.82 | 1.61 | 0.419 | 1.28 | 0.87 | 1.90 | 0.216 |
|  | g*BRCA2*pv DBD | 13 | 0.81 | 0.43 | 1.51 | 0.506 | 0.79 | 0.39 | 1.63 | 0.528 |
|  | g*BRCA2*pv outside domain | 35 | 2.10 | 1.46 | 3.00 | <0.001 | 2.03 | 1.44 | 2.87 | <0.001 |
|  | g*BRCA2*pv RAD51-BD | 39 | 1.37 | 1.01 | 1.85 | 0.04 | 1.58 | 1.14 | 2.21 | 0.006 |

DBD=DNA Binding Domain, RING=Really Interesting New Gene, RAD51-BD=RAD51 Binding Domain, BRCT=BRCA1 C-Terminal

Patients with *BRCA1* variants in exon 10 have been reported to have poorer outcomes^46^ due to expression of an alternative splice isoform called *BRCA1*-delta11q (Δ11q) that bypasses almost all of exon 10 of *BRCA1* (historically referred to as exon 11). To explore this further, we assessed *BRCA1* isoform expression in our multi-omics cohort (*n*=154) using the bulk RNA sequencing reads spanning the exon 10 to exon 11 junction (Fig. 2a, Supplementary Tables S7 and S8, Supplementary Information). The Δ11q isoform was widely expressed regardless of *BRCA-*status, but patients with *BRCA1* variants in exon 10 had significantly higher proportions of Δ11q transcripts relative to canonical transcripts (*P*=0.011; Fig. 2b). Patients with *BRCA1* variants in exon 10 were classified as having high (*n* = 10) or low (n = 9) *BRCA1* Δ11q expression, according to the median. Patients with high Δ11q expression had a shorter survival (median OS 2.74 years) compared to those with low Δ11q expression (median OS not reached), although this was not statistically significant (*P*=0.083) and was not associated with differences in the HRD sum score (Figs. 2c,d and Supplementary Table S9).

**Fig. 2.**
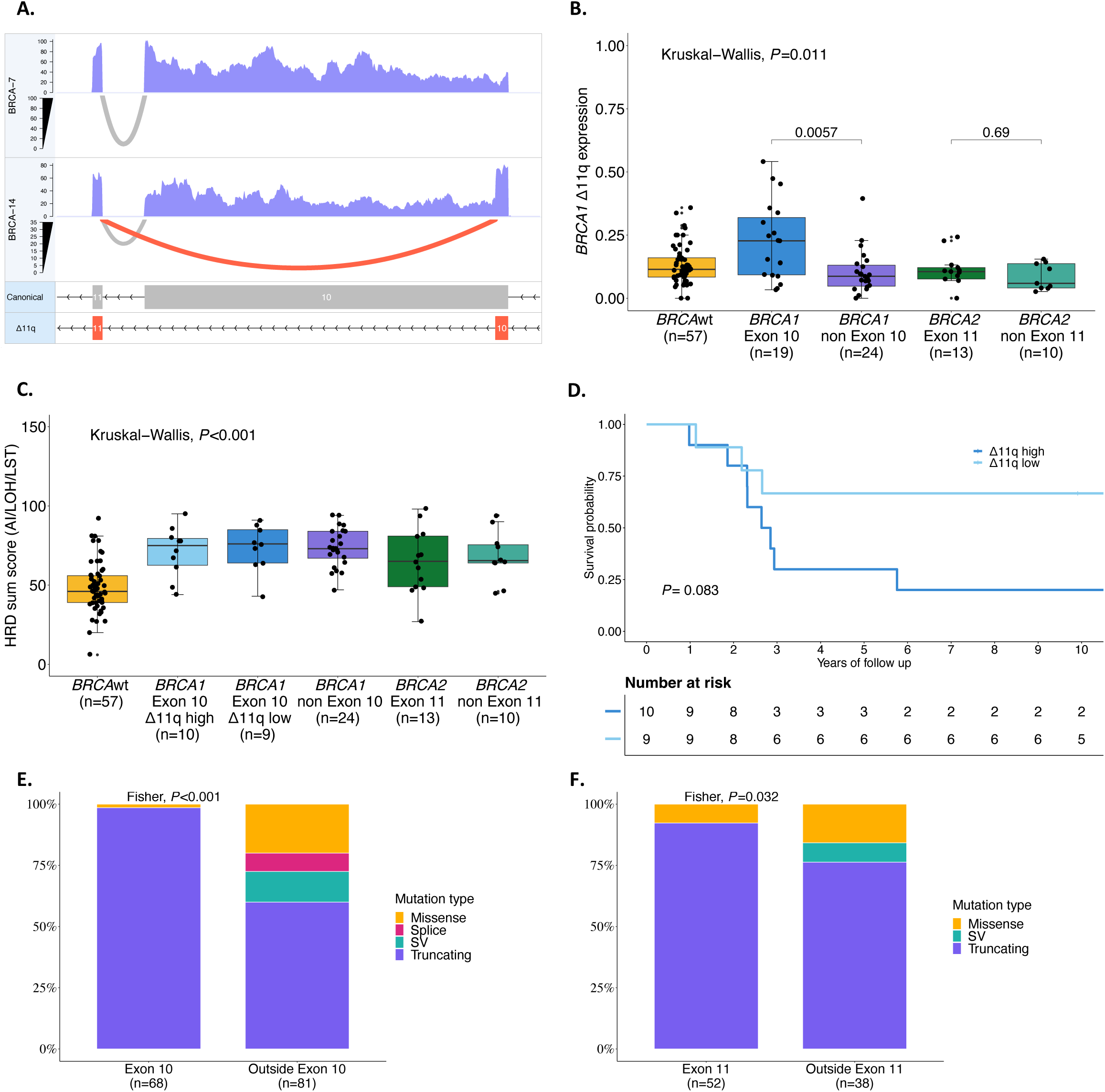
Analysis of pathogenic germline *BRCA1* and *BRCA2* variants and isoform expression on survival in HGSC. **a**, Illustrates the RNA-seq coverage and splice junction reads across the *BRCA1* gene for two samples (BRCA-7 and BRCA-14). The top and middle panels show the expression levels, with BRCA-7 and BRCA-14 indicating overall expression coverage. The bottom panel depicts the structure of the *BRCA1* isoforms, where the canonical isoform includes exon 10, while the Δ11q isoform excludes it. Grey arcs in the top and middle panels represent splice junction reads supporting the canonical isoform, while red arcs indicate reads supporting the Δ11q isoform. The higher expression of the Δ11q isoform in BRCA-14 compared to BRCA-7 highlights differential splicing events between these samples. **b**, Illustrates a comparison of *BRCA1* Δ11q expression among patients with mutations in *BRCA1* exon 10 and outside exon 10, *BRCA2* exon 11 and outside exon 11, and patients with *BRCA* wildtype. Kruskal–Wallis test *P* value is reported as well as pairwise Wilcoxon rank-sum test *P* values. **c**, Shows HRD sum score distribution among patients with mutations in *BRCA1* exon 10 (high and low Δ11q expression) and outside exon 10, *BRCA2* exon 11 and outside exon 11 and *BRCA* wildtype tumors. Kruskal–Wallis test *P* value is reported as well as pairwise Wilcoxon rank-sum test *P* values. **d**, Kaplan-Meier analysis of overall survival comparing high vs low Δ11q expression (divided by median) in patients with a *BRCA1* mutation on Exon 10. *P* value calculated by log-rank test. The distribution of mutation types within *BRCA1* outside exon 10 vs. on exon 10 and for *BRCA2* outside exon 11 vs. on exon 11 is presented in **e** and **f**, respectively. Fisher’s exact test *P* values are reported. *BRCAwt=BRCA wildtype, HR=Hazard ratio, n=Number of patients, SV= Structural variants, n= number of patients, LST=Large scale transitions, LOH= Loss of heterozygosity, AI= Allelic imbalance*

Overall, patients with g*BRCA2*pv had an improved OS compared to non-carriers, regardless of mutation location (Table 3). The only exception was the small group (*n*=13) with pathogenic variants in the DNA binding domain (DBD) of *BRCA2,* located outside of exon 11, who did not show a statistically significant OS or PFS benefit compared to non-carriers (TR: 0.79 and 0.81, 95% CI: 0.39-1.63 and 0.43-1.51, *P*=0.528 and *P*=0.506, respectively).

The type of mutation in *BRCA1* and *BRCA2* also plays a predictive role in response to PARPi therapy in ovarian cancer^43^. In our analysis, pathogenic variants in *BRCA1* exon 10 and *BRCA2* exon 11 were more likely to be truncating (98.6% and 92.3%) than those outside these exons (60% and 76.3%, *P*<0.001 and *P*=0.032 respectively; Figs. 2e,f). *BRCA1* and *BRCA2* domains associated with prolonged survival were more likely to have truncating variants than missense or splice site variants (*P*<0.001 and *P*=0.067, respectively; Extended Data Figs. 4c,d).

### *NF1* gene alterations are associated with improved survival in *BRCA2*-deficient HGSC

To identify genomic features associated with short survival in HRD tumors, we compared tumor genomes and transcriptomes between short (OS ≤3 years, STS) and long-term (OS >3 years, LTS) survival groups (Fig. 3a). Tumor genomes were classified as either *BRCA1*-deficient, *BRCA2*-deficient or *BRCA*-proficient, which incorporated germline and somatic alterations in *BRCA1* and *BRCA2*, as well as other well-defined HR genes, and tumor HRD status as determined by a mutational signature-based classifier (CHORD, Classifier of HOmologous Recombination Deficiency)^60^ (Supplementary Information and Supplementary Tables S10-S12). *CCNE1* amplifications (gene level copy number ≥7) were associated with *BRCA*-proficiency, and particularly the short-survival *BRCA-*proficient group (50%, *P*_adj_<0.001; Fig. 3b). *BRCA*-proficient tumors had less genomic scarring and were associated with an older age at diagnosis compared to *BRCA1*-deficient and *BRCA2*-deficient tumors (Extended Data Figs. 5a,b). Gene methylation has been identified as a prognostic factor in HGSC^61^, but no significantly differentially methylated genes with corresponding up- or down-regulated gene expression were observed between STS and LTS groups in *BRCA1*- and *BRCA2*-deficient tumors (Supplementary Table S13 and Supplementary Information).

**Fig. 3.**
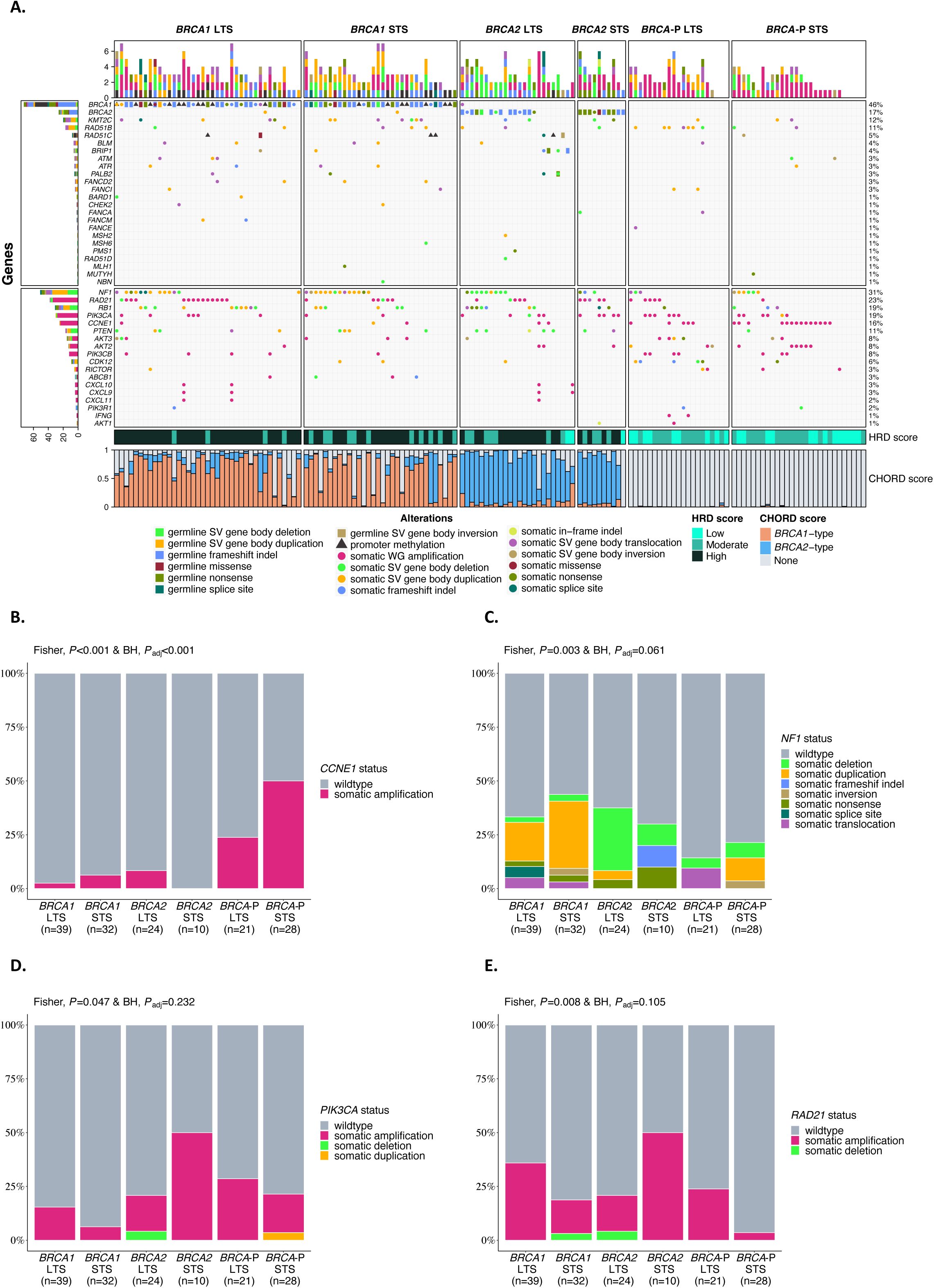
Genetic landscape of HGSC stratified by *BRCA* status and survival. **a**, Oncoprint showing germline and somatic alterations of homologous recombination (HR) genes and other genes of interest stratified by *BRCA*-status and survival group. The distribution of the mutation type within the *BRCA* survival group is shown for **b** *CCNE1*, **c** *NF1*, **d** *PIK3CA*, and **e** *RAD21. P*-values were calculated by the Fisher’s exact test and Benjamini-Hochberg (BH) adjusted (*P*_adj_). *BRCA status group: Long-term survivor (LTS) = OS >3 years, Short-term survivor (STS) = OS ≤3 years, BRCA-P=BRCA-proficient, HRD score: High= ≥ 63 HRD Sum, Moderate=42-62, Low= ≤41 HRD Sum, HRD= Homologous recombination deficiency, CHORD= Classifier of HOmologous Recombination Deficiency, SV=Structural variant, WG=Whole gene, BH=Benjamini-Hochberg*

Alterations in *NF1* were most common in *BRCA*-deficient tumors, regardless of survival group (*BRCA1* STS 43.8%, *BRCA1* LTS 33.3%, *BRCA2* STS 30%, *BRCA2* LTS 37.5%, *BRCA*-P STS 21.4%, *BRCA*-P LTS 14.3%, *P*_adj_=0.061; Fig. 3c and Supplementary Table S14). Notably, gene breakage caused by large-scale deletions was enriched in *BRCA2*-deficient tumors in the LTS group. We hypothesized that not all alteration types equivalently disrupt gene function. Indeed, only 54.2% (26/48) of *NF1* alterations showed a locus-specific loss of heterozygosity (LOH) suggesting a loss-of-function (Supplementary Table S14 and Supplementary Information). Concordantly, *NF1* mRNA expression varied in tumors according to the type of *NF1* alteration and was particularly depleted in those with locus-specific LOH (*P*<0.0001; Extended Data Fig. 6a). Patients with tumors that harbored loss-of-function *NF1* alterations showed an improved survival compared to non-loss-of-function *NF1* alterations (median OS 11.92 years vs 5.17 years, *P*=0.032; Extended Data Fig. 6b). In particular, the combination of both *BRCA2*-deficiency and loss-of-function *NF1* alteration (*n*=11) was associated with the best survival outcome (median OS 16.96 years), almost twice as long as those with *BRCA2*-deficient tumors with no loss-of-function *NF1* alteration (median OS 8.84 years; Extended Data Fig. 6c and Supplementary Table S9).

NF1 protein expression was assessed by IHC in a larger cohort enriched for long-term survivors (*n*=658; Extended Data Fig. 1). NF1 protein loss was observed in 13.37% (*n*=88/658) of patients and was associated with improved survival compared to retained NF1 expression (median OS 4.70 vs. 3.58 years, *P*=0.028; Extended Data Fig. 7a). Although there were few patients with NF1 protein loss and germline *BRCA1* (*n*=21) or *BRCA2* (*n*=6) pathogenic variants, NF1 loss was associated with better survival in g*BRCA2*pv-carriers (median OS 8.05 years NF1 loss vs. 5.72 years NF1 retained) but not in g*BRCA1*pv-carriers (median OS 4.74 years NF1 loss vs. 4.69 years NF1 retained; Extended Data Fig. 7b). NF1 loss also was associated with a longer survival among non-carriers (median OS 5.01 years NF1 loss vs. 3.36 years NF1 retained; Extended Data Fig. 7b).

In the independent OTTA cohort with *NF1* mRNA expression and survival data available (n=5666), low *NF1* expression (lowest quantile) was associated with improved survival compared to high expression (2^nd^ to 5^th^ quantiles) (median OS 4.19 vs. 3.56 years, *P*<0.0001; Extended Data Figs. 1 and 7c). Consistent with the other cohorts, g*BRCA2*pv-carriers with low *NF1* expression (n=36) showed an improved survival (median OS 6.42 years *NF1* low vs. 5.66 years *NF1* high), while there was no effect in g*BRCA1*pv-carriers (median OS 5.41 years *NF1* low vs. 5.65 years *NF1* high, Extended Data Fig. 7d).

### *PIK3CA* and *RAD21* amplifications are associated with short survival in *BRCA2*-deficient HGSC

We found an enrichment of *PIK3CA* and *RAD21* gene amplifications in *BRCA2*-deficient tumors in patients with short compared to long survival (*PIK3CA:* 5/10, 50% vs 4/24, 16.7%, *P*_adj_=0.232 and *RAD21:* 5/10, 50% vs 4/24, 16.7%, *P*_adj_=0.105, respectively; Fig. 3d,e). Co-occurrence of *RAD21* and *PIK3CA* amplification was observed in 8.8% (3/34) patients with *BRCA2*-deficiency (Supplementary Tables S15 and S16). *PIK3CA* and *RAD21* mRNA expression was highly correlated with copy number (*P*<0.0001), and tumors with gene amplification (≥7 copies) had a significantly higher expression (*P*<0.001 and *P*=0.02, respectively) (Extended Data Fig. 8a,b and Supplementary Tables S17 and S18). Patients with combined *BRCA2*-deficiency and *PIK3CA* amplification (*n*=9, median OS 2.89 years) or *RAD21* amplification (*n*=9, median OS 2.89 years) had a significantly worse prognosis compared to patients with *BRCA2*-deficient tumors without *PIK3CA* amplification (n=25, median OS 11.92 years) or *RAD21* amplification (*n*=25, median OS 11.53 years; Extended Data Fig. 8c,d and Supplementary Table S9).

PI-3 kinase pathway activity is thought to contribute to tolerance to genome doubling and *PIK3CA* amplification in whole-genome duplicated tumors is a frequent event in HRD end-stage HGSC^49,62^. The STS *BRCA2*-deficient group was characterized by high ploidy (*P*_adj_=0.0073) and whole-genome duplication (*P*_adj_=0.0404), in contrast to *BRCA1*-deficient and *BRCA*-proficient tumors where the LTS groups tended to have higher ploidy (Extended Data Fig. 5a). The association between *PIK3CA* and survival by *BRCA* status was further corroborated in the OTTA cohort, where g*BRCA2*pv carriers with high *PIK3CA* RNA expression (highest quantile) had shorter survival relative to their counterparts with low expression (median OS 4.09 vs 7.43 years, *P*<0.0001; Extended Data Fig. 8e). By contrast, g*BRCA1*pv carriers with high *PIK3CA* RNA expression showed improved survival (median OS 7.67 vs 5.23 years).

### Elevated HRD scarring is prognostic for survival in *BRCA*-deficient HGSC

High tumor mutation burden has been shown to be associated with long-term survival in ovarian cancer^22^. However, we found that tumor mutation burden and predicted neoantigen counts were equivalent in *BRCA1*-deficient and *BRCA2*-deficient tumors between STS and LTS groups (Fig. 4a-c, Extended Data Fig. 5a, and Supplementary Table S19). Among various genomic features that were compared between these groups (Extended Data Fig. 5a), the HRD score^27^ was elevated in *BRCA1*-deficient tumors with long survival times compared to those with short survival times (*P*=0.017; Fig. 4d). HRD score is a measure of genomic scarring associated with impaired HR repair, suggesting a more profound inactivation of the HR pathway in patients with good outcome. Retention of the wildtype allele with absence of locus specific LOH has been reported to influence outcomes in g*BRCA*pv-carriers in ovarian and breast cancer^63–66^. However, in our cohort there was only one g*BRCA2*pv carrier without loss of the wildtype allele (patient BRCA_9; Supplementary Table S11 and Supplementary Information). Concordantly this tumor was HR-proficient with an HRD score of 27 (HRP ≤42 HRD sum score) and CHORD score of 0 (HRP ≤0.5 CHORD score), and the patient had short OS (<3 years).

**Fig. 4.**
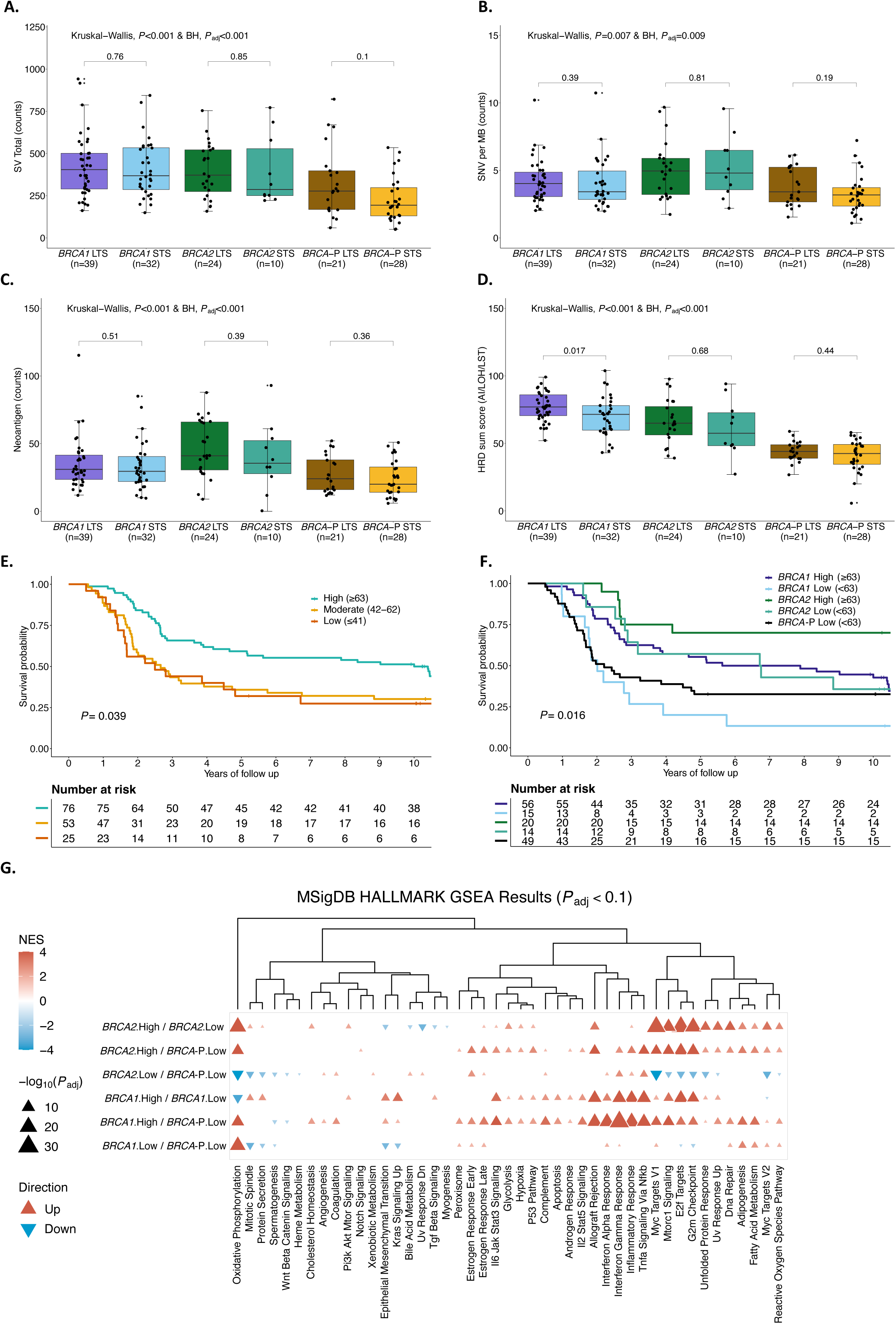
Influence of homologous recombination deficiency in HGSC independent of *BRCA* status. Comparison of **a** SV total counts, **b** SNV counts per megabase, **c** neoantigen counts, and **d** HRD sum score between *BRCA* survival groups (*BRCA1=BRCA1*-deficient; *BRCA2=BRCA2*-deficient; *BRCA-P*=*BRCA*-proficient; Long term survivor (LTS) = OS >3 years; Short term survivor (STS) = OS ≤3 years). *P*-values were calculated by the Kruskal Wallis test and Benjamini-Hochberg (BH) adjusted (*P*_adj_). **e**, Kaplan-Meier analysis of overall survival stratified by different thresholds of the HRD sum score (High ≥63, Moderate 42-62, Low ≤42) in 154 patients with *BRCA*-deficient and *BRCA*-proficient HGSC. *P* value calculated by log-rank test. **f**, Kaplan-Meier analysis of overall survival in patients with HGSC stratified by *BRCA*-status and high (High ≥63) or low (Low < 63) HRD sum score. *P* value calculated by log-rank test. **g**, Clustered heatmap summarizing gene set enrichment analysis (GSEA) using the hallmark Molecular Signatures Database (MSigDB) gene sets. Direction and color of triangles relate to the normalized enrichment score (NES) as generated by FGSEA. P values (two-sided) were calculated using the FGSEA default Monte Carlo method; the size of the triangles corresponds to the negative log10 Benjamini-Hochberg (BH) adjusted P value (*P*_adj_). Columns are separated by *BRCA-*status and HRD score groups (*BRCA1*; *BRCA2*; *BRCA*-P, *BRCA*-proficient, High ≥ 63; Low <63) with the direction of enrichment indicated by the first group mentioned in the x-axis label. *SV=Structural variants, SNV=Single nucleotide variant, MB=Megabase, HRD=Homologous recombination deficiency, HRP=Homologous recombination proficiency, BRCA-P=BRCA-proficient, LST=Large scale transitions, LOH= Loss of heterozygosity, AI= Allelic imbalance*

We observed a dynamic range in HRD scores, even among tumors with pathogenic *BRCA* mutations, suggesting a non-equivalence of alterations. The cutoff of the HRD score has been debated, with 42 mainly used in recent clinical trials^67–71^, and a more stringent threshold of 63 has been proposed for ovarian cancer^72^. Indeed, patients whose tumors had a high HRD score (≥63) had longer OS (median OS 10 years) compared to those with HRD scores of 42-62 (median OS 2.66 years) and ≤41 (median OS 2.5 years), regardless of *BRCA*-status (*P*=0.039; Fig. 4e and Supplementary Table S9). Applying a threshold of 63 to divide samples into high and low HRD, all *BRCA*-proficient tumors had a low HRD score. Furthermore, patients with *BRCA1*- and *BRCA2*-deficient tumors and HRD scores ≥63 had longer OS compared to patients with lower HRD scores (median OS 6.76 vs. 2.01 years and 11.88 vs. 6.73 years, respectively; Fig. 4f and Supplementary Table S9). Notably, patients with *BRCA1*-deficient tumors with HRD scores <63 had similar OS to patients with *BRCA*-proficient tumors (median OS 2.01 years vs 2.21 years).

Gene set enrichment analysis^73^ (GSEA; Methods) revealed distinct patterns of pathway regulation based on HRD scores and *BRCA* status in patients with HGSC. Specifically, pathway activation in *BRCA1*- and *BRCA2*-deficient patients with low HRD (<63) closely resembled those of *BRCA*-proficient patients (Fig. 4g). In contrast, *BRCA1*-deficient patients with high (≥63) HRD scores showed an upregulation of several pathways, including interferon-gamma and inflammatory response. These pathways are primarily involved in host defense and immune surveillance^74^, underscoring their potential role in modulating the tumor microenvironment and influencing immune response in patients with *BRCA1*-deficient tumors.

### CD8+ PD-1+ T cells are prognostic for survival in g*BRCA*pv-carriers

We considered whether *BRCA*-deficient cases with shorter survival would have fewer mutation-associated neoantigens to drive anti-tumor responses, but there was no difference in neoantigen counts between the STS and LTS groups for both *BRCA1* and *BRCA2* (*P*=0.51 and *P*=0.39, respectively; Fig. 4c). Tumor samples from 143 HGSC g*BRCA*pv-carriers were analyzed by multi-color immunofluorescence to determine the epithelial and stromal immune cell densities and their associations with survival groups (Extended Data Fig. 1). Aside from intraepithelial B cells and CD4+ T cells (OR=1.0), all other immune cell subsets had a positive association with survival (OR<1.0; Supplementary Table S20). Only intrastromal and intraepithelial CD8+ PD-1+ T cells were significantly more abundant in g*BRCA*pv-carriers with LTS compared to those with STS (*P*=0.043 and *P*=0.029, respectively; Supplementary Table S20).

### The mesenchymal features *c-KIT* and mast cells are associated with poor outcome in HGSC

Immune cell abundance was estimated in 154 HGSC tumor samples using CIBERSORTx^75^. Unsupervised clustering of the inferred immune cell densities identified six groups of patients (Fig. 5a, and Supplementary Table S21) associated with differential survival outcomes (*P*=0.0053; Fig. 5b). The IMMB.1 (*n*=30) and IMMB.6 (*n*=25) clusters had exceptionally long survival (median OS 14.87 and 10.45 years, respectively; Supplementary Table S9). The group with the shortest survival (cluster IMMB.5, *n*=24, median OS 2.03 years) was enriched with activated dendritic cells and resting mast cells, a feature associated with the C1.MES subtype (*P*=0.0021; Fig. 5c). Multivariable Cox regression analysis showed that resting mast cells (HR: 1.26, 95% CI 1.06-1.5, *P*=0.009) were the immune cell type most strongly associated with short survival (Extended Data Fig. 9a). *BRCA1*-deficient tumors in patients with STS had increased expression of the mast cell growth factor receptor *c-KIT* (CD117) compared to those with LTS (*P*=0.003, *P*_adj_=0.101; Extended Data Fig. 9b). Patients with high *c-KIT* tumor expression had significantly shorter OS than those with low *c-KIT* tumor expression, regardless of *BRCA* and HRD status (HR: 1.71, 95% CI 1.16-2.53, *P*=0.0071; Extended Data Fig. 9c). The C1.MES subtype showed higher expression of *c-KIT*, together with an upregulation of the epithelial mesenchymal transition (EMT) pathway, compared to the C2.IMM subtype (*P*_adj_<0.001) (Extended Data Fig. 9d,e).

**Fig. 5.**
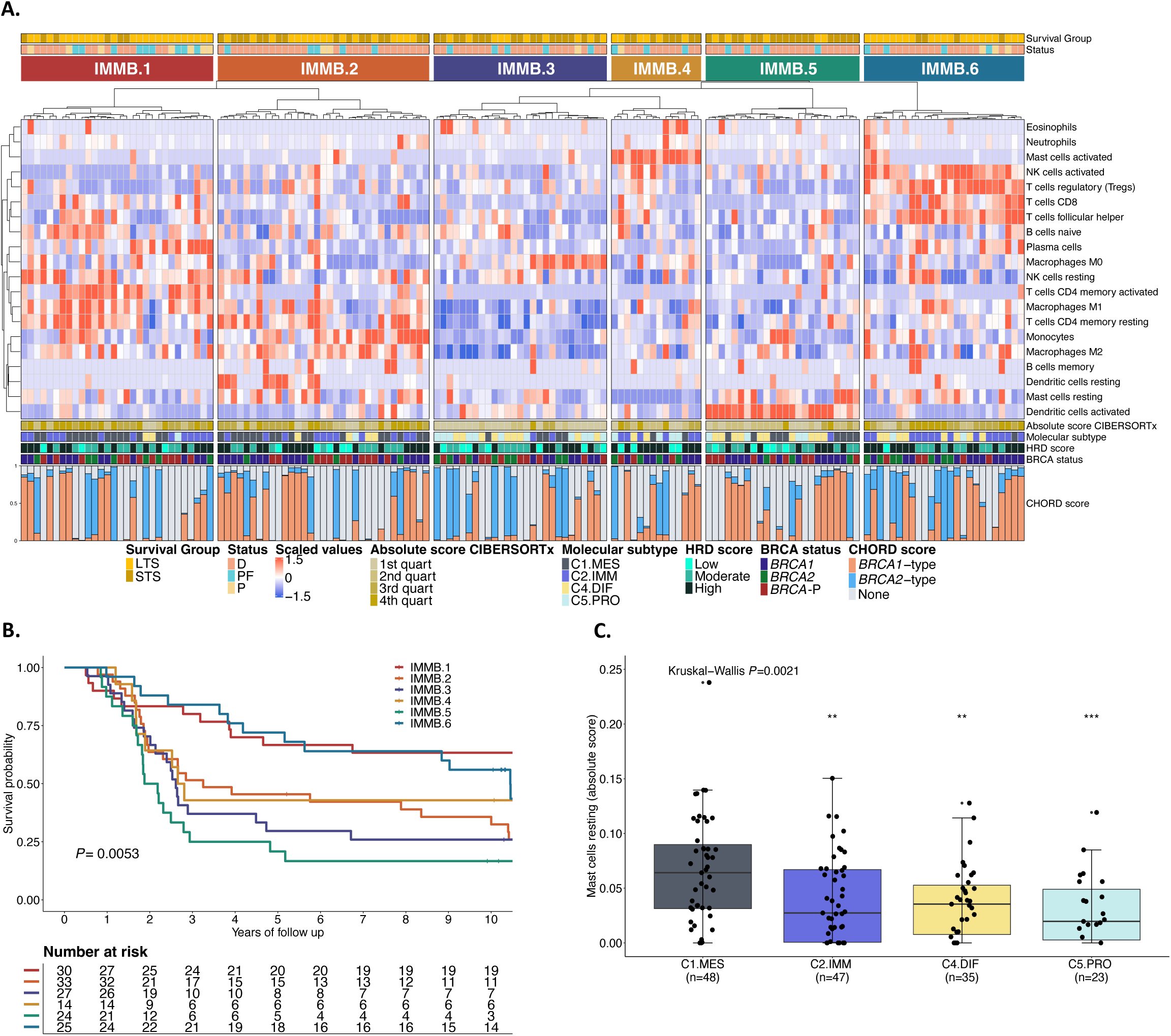
Integration of immune cell profiling by CIBERSORTx and survival analysis in HGSC. **a**, Summary of the immune cell types arising from the CIBERSORTx analysis from *BRCA-*deficient and *BRCA*-proficient samples (n = 153 patients). Tumors fell into 6 major clusters (IMM.1-IMM.6) of immune cell types associated with survival. Each patient is annotated with survival group, status at last follow-up, CIBERSORTx absolute immune scores, molecular subtype, HRD score, *BRCA* status and CHORD score. **b**, Kaplan-Meier analysis of overall survival stratified by immune clusters. *P* value calculated by log-rank test. **c**, Boxplots summarize the absolute cell enrichment score of mast cells resting markers across the molecular subtype (C1.MES; C2.IMM; C4.DIF; C5.PRO); points represent each sample, boxes show the interquartile range (25–75th percentiles), central lines indicate the median, and whiskers show the smallest/largest values within 1.5 times the interquartile range. Kruskal– Wallis test *P* value is reported as well as pairwise Wilcoxon rank-sum test *P* values comparing molecular subtypes (C2.IMM; C4.DIF; C5.PRO) to C1.MES (**, *P* <0.01, ***, *P* <0.001). *Survival group: Long term survivor (LTS)= OS >3 years, Short term survivor (STS)= OS ≤3 years, HRD=Homologous recombination deficiency, HRD score: High= ≥ 63 HRD Sum, Moderate=42-62, Low= ≤41 HRD Sum, Molecular subtypes: C1.MES=C1 mesenchymal subtype, C2.IMM=C2 immunoreactive subtype, C4.DIF=C4 differentiated subtype, C5.PRO=C5 proliferative subtype, Status: D=Dead, PF=Progression-free, P=Progression, IMMB=Immune cluster BadBRCA (IMMB.1-IMMB.6),*

## DISCUSSION

Our study highlights the complexity of survival determinants in patients with HGSC, demonstrating that it is the intersection of multiple factors, including surgical residual disease, immune response, and somatic gene alterations, which may influence outcome rather than *BRCA* mutation status alone. This interplay was particularly apparent in the diminished adverse impact of surgical residual disease in g*BRCA*pv-carriers compared to non-carriers. Previous reports have suggested that surgery in a *BRCA*-deficient setting may have a lesser impact on survival in both first-line and platinum-sensitive setting^33,47,76^, indicating that it may be particularly important to achieve complete resection of *BRCA*-proficient tumors. In addition, an exploratory analysis of the PAOLA-1/ENGOT-ov25 trial^77^ showed that patients with *BRCA*-proficient tumors classified as higher risk (FIGO stage III with primary cytoreductive surgery and residual disease, or NACT; FIGO stage IV) had notably worse PFS compared to lower-risk patients, while this difference was less pronounced in patients with *BRCA*-deficient tumors. These results emphasize the importance of primary cytoreductive surgery with complete resection for non-carriers, who may also benefit more from secondary cytoreductive surgery in contrast to g*BRCA*pv-carriers^78^. Equally, it may be that the positive effect of optimal cytoreduction is not as apparent in *BRCA* carriers, due to the chemotherapy (platinum) sensitivity associated with *BRCA*-deficiency.

In the current study, the association between NACT and survival appeared to differ by g*BRCA*pv status, with a potential attenuation of survival benefit among g*BRCA*pv-carriers who received NACT. However, the subgroup analyses by g*BRCA*pv status and treatment type were likely underpowered, limiting definitive conclusions regarding potential interactions. Given the rapid increase in the uptake of NACT in recent years^79^, it will be important to determine if patients with *BRCA*-deficient tumors may be negatively impacted by NACT^80^. The acquisition of *BRCA* reversion mutations is frequent^48–50^, and it is plausible that reversion events may be more common where chemotherapy commences with a large tumor volume from which resistant clones could emerge under selection^58^. This is especially important in the PARPi era, where the early development of platinum resistance could negatively impact on the potential benefit gained from PARPi treatment. While the impact of NACT on outcomes according to *BRCA* status is not yet known, it is becoming increasingly important to more rapidly determine the *BRCA* and broader HR status of a patient’s tumor at diagnosis to make the most informed decisions at primary treatment.

Our study highlights the spectrum of HRD scores seen in patients with *BRCA*-deficient tumors. While all but two exceeded a threshold (>42) required for classification as HRD, the improved OS and PFS seen with a more stringent threshold (≥63) shows that HRD should not be considered a binary classification but rather appears to be a continuous variable. This finding is consistent with a previous analysis of 537 HGSC cases from The Cancer Genome Atlas which showed that patients with HRD scores ≥63 were associated with better survival outcomes, while those with intermediate (42-62) and low (≤42) HRD scores had overlapping survival curves^72^. It is important to mention that in our study, samples were collected over nearly 20 years, a timeframe that encompasses changes in treatment practices, making it challenging to determine how evolving therapies, particularly the introduction of PARPi, may have influenced outcomes. It is notable that the HRD score threshold of 42 was originally established to predict response to neoadjuvant platinum-containing chemotherapy in patients with breast cancer^81^, which tends to have less genomic scarring compared to ovarian cancer^27,72^. As HRD scores ≥63 strongly predicted better outcomes in *BRCA*-deficient HGSC, our findings support the prognostic value of HRD score thresholds. However, it is premature to conclude that a higher threshold should alter therapy selection. To establish this, a comprehensive analysis of maintenance PARPi trials, incorporating HRD scores, would be necessary to confirm their predictive role in guiding treatment decisions. Furthermore, it would be ideal to extend this investigation to include other relevant genomic alterations identified in trial samples to refine patient stratification further. This refinement would help identify patients for whom no maintenance therapy or additional targeted therapy may be more appropriate, while avoiding potentially ineffective treatments for those with lower HRD scores, thereby personalizing therapy to maximize efficacy and minimize unnecessary side effects.

Our analyses corroborated Labidi-Galy et al.’s findings that pathogenic variants in the RAD51-BD of *BRCA2*^44^ and the DBD of *BRCA1*^43^ are associated with improved outcomes in HGSC. By contrast, alterations outside *BRCA1* exon 10, particularly in the BRCT and RING regions, are not associated with a significantly improved survival compared to non-carriers and in some cases may confer platinum and PARPi resistance^45^. While *BRCA1* exon 10 mutations have been associated with improved outcomes in multiple studies, including ours, there is evidence that tumors may express the *BRCA1*-Δ11q splice isoform, which bypasses exon 10 mutations and results in a shorter but partially functional protein that is permissive of treatment resistance^43,46^. In a relatively small sample size for which we had RNA-seq data (n=19 *BRCA1* exon 10 mutated tumors), we found that patients with a pathogenic *BRCA1* variant in exon 10 and high Δ11q expression had a shorter survival. We were unable to measure Δ11q expression during or following treatment. This is important because Δ11q expression may increase or fluctuate under the selective pressure of treatment, which would influence treatment response and survival outcomes.

CD8+ PD1+ T cells are associated with improved outcomes in ovarian cancer^82^, contributing to enhanced anti-tumor immunity. In our analysis, the presence of these cells in tumors were prognostic for survival in g*BRCA*pv-carriers, although to a lesser extent. This suggests that while cytotoxic T-cell activity remains important in *BRCA*-deficient tumors, additional factors may influence survival. Given the established association between *BRCA* and HR status and increased TMB^22^, it is possible that immune exhaustion, suppressive signaling or tumor-intrinsic immune resistance pathways may counteract the expected immunogenicity. Intriguingly, *BRCA1*-deficient tumors with high HRD scores had evidence of enhanced immune-related gene transcription. In addition, while our study did not include cigarette smoking in the survival models, smoking has been identified as a potential factor influencing survival in g*BRCA*pv-carriers^83^, which may also influence the immune response. Further research into markers of T-cell exhaustion and other immune regulators is needed to better understand the differential immune responses in these patients.

NF1 gene loss-of-function emerged as a good prognostic factor in *BRCA2*-deficient HGSC. Loss-of-function of *NF1* is common in epithelial ovarian cancer with a prevalence of 12–31%^13,20,22,58,84,85^. NF1 inactivation by gene breakage or mutations may contribute to initial good prognosis but later chemoresistance in patients with HGSC and *BRCA*-deficiency^84^. This is consistent with recent findings that deleterious *NF1* mutations are associated with improved PFS in ovarian cancer^20^ and low mRNA expression of *NF1* predicts longer overall survival^22^. In contrast, *PIK3CA* amplification and high mRNA expression were associated with shorter survival in patients with *BRCA2*-deficient HGSC. As a major regulator of the phosphoinositide 3-kinase (PI3K) pathway, *PIK3CA* activation promotes cell proliferation and survival, especially in genomically unstable cancers^49,62^. Its amplification may enhance tolerance to genome doubling and contribute to the aggressive nature of *BRCA2*-deficient tumors. The contrasting survival outcomes between *PIK3CA* amplification and *NF1* loss-of-function underscore the heterogeneity of HGSC tumors, highlighting the need for personalized therapeutic strategies, even within the *BRCA2*-deficient subgroup.

## METHODS

### Ethics statement

Written informed consent or an approved waiver of consent was obtained at each participating study site for patient recruitment and the use of samples and linked clinical information (Supplementary Table S22). Investigations were performed after approval by local human research ethics/institutional review board committees at each site. This study was conducted in accordance with the principles of Good Clinical Practice, the Declaration of Helsinki and local regulations.

### Study population

This retrospective, multi-center study included patients diagnosed with HGSC between 2002 and 2019. The Australian Ovarian Cancer Study (AOCS) cohort (n=1389) included all stages (FIGO I-IV), and the Multidisciplinary Ovarian Cancer Outcomes Group (MOCOG) cohort (n=154) was restricted to advanced stage disease (FIGO III and IV; Table 1, Extended Data Fig. 1, and Supplementary Table S22). Patients were categorized based on OS into short (<3 years) and long (≥3 years) OS groups (Supplementary Information). For multi-omics analysis, 154 patients had fresh-frozen tumor obtained during primary cytoreductive surgery and matched blood samples, or were previously analyzed^22,58^. Findings were validated in an independent HGSC cohort (n=5875) from the Ovarian Tumor Tissue Analysis Consortium (OTTA) for which g*BRCA*pv status was available.

### Molecular data

#### Single-nucleotide polymorphism (SNP) arrays

Tumor and matched normal DNA was analyzed with the Infinium OmniExpress-24 BeadChip arrays as described previously^22^. The concordance of normal and tumor DNA was assessed using HYSYS^86^. Tumor DNA samples with estimated tumor cellularity >40% (determined by qPure^87^ and ASCAT^88^) were considered appropriate for whole genome sequencing and methylation arrays.

#### Whole genome sequencing (WGS)

For WGS, libraries were generated from tumor and matched normal genomic DNA from peripheral blood mononuclear cells with a minimum base coverage of 60x and 30x, respectively. FASTQ files were assessed for sequencing quality using FASTQC (v0.11.8) and, for contaminants using FastQ Screen^89^ (v0.11.4). Adapters, N-content and low-quality bases were trimmed using fastq-mcf (v1.05). Sequenced data was mapped to the human genome reference GRCh37 b37 using the aligner BWA mem^90^ (v.0.7.17-r1188). Aligned BAM files per lane were then sorted, merged and duplicates marked using Picard Tools (v.2.17.3). Further processing of the aligned files included base recalibration using GATK BaseRecalibrator (v4.0.10.1). Coverage calculation was performed using GATK DepthOfCoverage (v3.8-1-0-gf15c1c3ef). GATK HaplotypeCaller (v.4.0.10.1) was used on germline BAMs to generate Genomic Variant Call Format (GVCF) files which were used as the Panel of Normals (PoN) in the Mutect2 somatic variant calling workflow. Tumor purity and ploidy were estimated using FACETS^91^.

#### RNA-sequencing (RNA-seq)

Extracted RNA from tumor tissue samples underwent RNA-seq, with initial quality control checks on raw FASTQ files performed using FastQC^89^ (v0.11.8). Adapter, poly (A) tails, N content and low quality base trimming was done using fastq-mcf (v1.05), and contamination was assessed using FastQ Screen^89^ (v(0.11.4). Reads were then mapped to the human reference GRCh37.92 using the STAR^92^ (v2.6.0b) two-pass method. The mapped reads were then sorted using Picard Tools (v2.17.3). Counts were generated using HTSeq^93^ (v0.10.0) on the GRCh37.92 Ensembl release gene annotation. Raw count data was then subsetted to protein coding genes and lowly expressed genes were removed using the following strategy. First, raw counts were converted to CPM (counts per million) and only protein coding genes with a CPM of greater than 0.5 in at least 10 samples were retained. The resulting raw count matrix was then normalized using the trimmed mean of M values (TMM) method using edgeR^94^ (v3.28.1). Batch effects were removed using limma’s^95^ (v3.48.2) removeBatchEffect function. Batch effect removal was done by applying batch correction on the library type (stranded/unstranded) while preserving the survival group (long/short).

#### Methylation arrays

The generation and processing of methylation array data was performed as previously described by Garsed et al.^22^. Briefly, initial quality control was performed by QuantiFluor (Promega). Subsequently, 500 ng tumor DNA was converted using the EZ DNA Methylation kit (Zymo Research) and analyzed using the Infinium MethylationEPIC BeadChip arrays. The R package minfi^96^ (v1.32.0) was then used for quality control assessment and processing of the methylation data as previously described^22^.

#### Immunofluorescence (IF) data

Tissue microarrays (TMAs) were constructed from formalin-fixed paraffin-embedded (FFPE) blocks of tumor tissue and stained by IF with two panels of antibodies against immune markers of interest. Panel 1 detected CD3, CD8, CD20, FOXP3 and CD79; panel 2 detected CD3, CD8, PD-1, PD-L1 and CD68. Both panels also detected pan-cytokeratin to identify tumor epithelium. Automated cell scoring, including separation of epithelial and stromal regions, was performed using QuPath (v0.2m2), with extensive manual training and validation. CD4+ T cells were defined as CD3+CD8-cells, as previously^97^.

#### Immunohistochemistry (IHC)

Sections of 4 µm thickness were cut from previously constructed TMAs of FFPE tumor samples. Deparaffinized sections were stained with the C-terminal NF1 antibody (clone NFC, SIGMA #MABE1820; St. Louis, MO, USA) using our previously described protocol on a DAKO Omnis platform: 30 min of pre-treatment heat-induced antigen retrieval in Tris-EDTA buffer, pH = 9.0; primary antibody incubation for 1h at dilution 1/50, 10 min of a mouse linker, and 30 min for the peroxidase labelled Dako EnVision+polymer-based detection system (Dako protocol 1 h-10M-30, Agilent, Santa Clara, CA, USA)^85^. Samples were scored as follows: inactivated (loss of expression with retained internal control), normal retained expression, subclonal loss, uninterpretable (loss of tumor expression but no internal control present), and exclude (no tumor in core) (**Supplementary Information**).

#### mRNA expression data by NanoString

Tumor mRNA expression data for genes of interest (*NF1, PIK3CA, c-KIT,* and *RB1*) and transcriptional molecular subtypes in the OTTA cohort were determined using NanoString, as previously described^98,99^.

### Measurements

#### Variant detection and annotation

Variant calling was performed for:

1. germline base substitution and INDEL variants by VarDictJava^100^ (v1.5.7 with –r = 2 –Q = 10 –f = 0.1).
2. somatic base substitution and INDEL variants using four separate variant callers as follows: by Mutect2^101^ (v4.0.11.0 with defaults), VarDictJava^100^ (v1.5.7 with –r = 2 –Q = 10 –V = 0.05 –f = 0.01), Strelka2^102^ (v2.9.9 with defaults), and VarScan2^103^ (SAMtools^104^) v1.9 for mpileup and VarScan2 v2.4.3 with -min-coverage 7 -min-var-freq = 0.05 -min-freq-for-hom = 0.75 -p-value = 0.99 -somatic-p-value =0.05 -strand-filter =0). Variant calls were decomposed and normalized using vt^105^ GATKs ReadBackPhasing tool (v3.8-1-0-gf15c1c3ef with -phaseQualityThresh = 10 – enableMergePhasedSegregatingPolymorphismsToMNP -min_base_quality_score =10 - min_mapping_quality_score = 10 -maxGenomicDistanceForMNP = 2) was applied on the passing variants per tool to combine contiguous SNVs to MNVs (multi-nucleotide variants). GATK’s CombineVariants (v3.8-1-0-gf15c1c3ef with -genotypeMergeOptions UNIQUIFY -priority Strelka2, Mutect2, VarScan2, VarDictJava) was used to merge the variant calls from all four callers into a consensus variant call set. The resulting variant call format (VCF) file was once again decomposed and normalized using vt. Forward and reverse strand counts for the reference and alternate alleles were calculated using bam-readcount (v0.8.0). Finally, all variants were annotated for Duke and DAC blacklisted regions. Any variants that were passed in at least two callers, had at least one variant read in each strand, and were not in the database of FrequentLy mutAted GeneS (FLAGS)^106^ or the Duke and DAC blacklist regions were deemed high-confidence.
3. structural variants (SV) using four separate callers Manta^107^ + BreakPointInspector (v1.5.0), GRIDSS^108^ (v2.0.1), Smoove (v0.2.2) and SvABA^109^ (v134). The SV calls were split into germline and somatic VCFs per caller. The findBreakpointsOverlaps method of the R library StructuralVariantAnnotation (v1.3.1) with a value of 10 for the ‘maxgap’ parameter was used to intersect common breakpoints between the callers. SVs were annotated to constituent types (duplication, deletion, inversion or translocation) using a simple annotation script provided by the GRIDSS tool. High-confidence SVs were categorized as those called by two or more callers.
4. copy number variations (CNV) detection by FACETS^91^ and cnv_facets (v0.13.0) as described previously^22^.

The detected variants were filtered for variants with a high probability of pathogenicity as described in detail before^22^.

#### Mutation burden and downsampling

We downsampled the higher coverage tumor BAM files using Picard DownsampleSam (v2.17.3) to achieve balanced median coverage sequencing batches, to compare mutation burden across samples with inconsistent coverage^22^. The median coverage of the International Cancer Genome Consortium (ICGC) tumors was 52.15x, the MOCOG tumors was 77.81x and the short survival *BRCA* dataset tumors was 64.98x. So, to get the same median coverage across the three batches we downsampled the MOCOG and short survival *BRCA* dataset tumors to the ICGC median by specifying downsampling fractions of 0.67 and 0.8 respectively. See **Supplementary Table S19** for details on the tumor sample coverage before and after downsampling and the number of SNVs, MNVs, indels and SVs called after downsampling.

#### Neoantigen prediction

Neoantigen prediction was performed as previously reported by Garsed et al.^22^. Briefly, HLA-VBSeq^110^ (v11_22_2018) was used to generate HLA types which were then used to identify and construct neoantigen using pVACtools^111^ pVACseq (v1.3.5).

#### Homologous recombination deficiency (HRD)

HRD status was determined using 1) scarHRD^112^, which uses loss of heterozygosity (LOH), telomeric allelic imbalance (TAI), and large scale state transition (LST) in tumor genomes to generate a HRD sum score, and 2) CHORD (Classifier of Homologous Recombination Deficiency)^60^, which uses specific base substitution, indel and structural rearrangement signatures detected in tumor genomes to generate *BRCA1*-type and *BRCA2*-type HRD scores.

#### RNA-seq data analysis

Raw count data was subsetted to protein coding genes and lowly expressed genes were removed using the following strategy. First, raw counts were converted to CPM (counts per million) and only protein coding genes with a CPM of greater than 0.5 in at least 10 samples were retained. The resulting raw count matrix was then normalized using the trimmed mean of M values (TMM) method using edgeR^94^ (v3.28.1). Batch effects were removed using limma’s^95^ (v3.48.2) removeBatchEffect function. Batch effect removal was done by applying batch correction on the library type (stranded/unstranded) while preserving the survival group (long-term/short-term).

#### RNA differential expression and pathway analysis by grouping

##### Groupings

For differential expression and pathway analysis, various groupings were used alone or in combination, namely 1) *BRCA*-deficiency status, 2) HRD groups, survival groups, and 3) molecular subtypes (**Supplementary Information**).

##### Differential expression analysis

To identify differentially expressed protein-coding genes between the comparison groups of interest, DESeq2 (v1.26.0)^113^ was applied. Raw counts were filtered to remove low expressed genes prior to analysis and batch effects were accounted for in the model^22^.

##### Gene Set Enrichment Analysis (GSEA)

FGSEA v1.15.1 was used to calculate gene set enrichment across the comparison groups. *P*-values obtained from DESeq2 were transformed to signed *P*-values and then sorted and fed into FGSEA to generate enrichment scores and FDR-adjusted *P*-values across the Hallmark gene sets in the MSigDB database49 (v7.4) via its function fgseaMultilevel (minSize=15, maxSize = 500, gseaParam = 0, eps = 0)^22^.

#### CIBERSORTx

CIBERSORTx analysis was performed as previously described^22^. Briefly, CIBERSORTx^75^ with the LM22 matrix was used on RNA-seq data for immune cell deconvolution. Immune clusters were then generated with k-means clustering of the generated absolute cell abundances using ConsensusClusterPlus^114^ (Supplementary Information).

#### Immunofluorescence

Data were categorized based on epithelial content, measured directly by pan-cytokeratin positivity and cell morphology (assessed by automated image analysis). Epithelium-negative, cellular (i.e., non-necrotic) tumor regions were defined as stroma. Immunomarker density (D; cells/mm^2^) for a given marker was calculated separately for epithelial and stromal compartments. For cases with multiple cores, the epithelial area was taken as the sum of all their individual TMA epithelial areas and similarly for the stromal area. We categorized marker D values into quartiles (separately for epithelial and stromal markers) to provide categorical comparisons for ease of interpretation of the odds ratios (ORs). Conditional logistic regression models were fitted for the long survival group vs short survival group. Logistic regression analyses were performed with the quartile values (scored as 1, 2, 3, 4). Immune clusters were then generated by k-means clustering of the immune cell type densities using ConsensusClusterPlus^114^.

### Statistical analyses

Continuous variables were compared between groups using the Kruskal-Wallis test and the difference between proportions of categorical data were assessed using the Chi-squared or Fisher’s exact test. Correlations between continuous variables were assessed using Spearman correlation. Benjamini-Hochberg adjusted *P*-values are reported as *P*_adj_ to account for multiple testing. Median PFS and OS were estimated using the Kaplan-Meier method and survival distribution were compared using the log-rank (Mantel-Cox) test.

For the AOCS cohort, univariable and multivariable survival analyses were performed using Accelerated Failure Time (AFT) models^54^ with a log-logistic distribution to evaluate associations between clinical and molecular variables and time-to-event outcomes. Results were reported as Time Ratios (TR) with 95% confidence intervals (CI), where a TR > 1 indicates longer time to progression or death, and a TR < 1 indicates shorter survival. Wald tests were used to compute *P*-values for individual covariates and interaction terms. Age at diagnosis was modelled using restricted cubic splines with three knots to allow for potential non-linear effects. Model assumptions were assessed using quantile-quantile plots of deviance residuals and Cox-Snell residuals to evaluate overall model fit. The Akaike Information Criterion (AIC) was used to compare alternative parametric distributions and confirm the suitability of the log-logistic model^115^.

For survival analyses of the OTTA cohort, Cox proportional hazards models were applied. Left truncation was used to account for delayed study enrolment at some sites, and follow-up time was right-censored at 10 years from diagnosis to minimize the influence of non-ovarian cancer-related deaths. *P*-values from Cox models correspond to Wald and log-rank tests. The proportional hazards assumption was assessed using the Grambsch-Therneau test based on scaled Schoenfeld residuals and further evaluated through graphical inspection of Schoenfeld residual plots^115,116^.

All statistical tests were two sided and considered significant when *P* < 0.05 or *P_adj_* <0.1. All analyses were performed using the statistical software R version 4.1.3^117^.

## Data availability

### Short survival BRCA dataset

WGS, RNA-seq and SNP array data from short-term survivors generated as part of the current study have been deposited in the European Genome-phenome Archive (EGA) repository (https://ega-archive.org) under accession code EGAS00001008059. WGS and RNA-seq data are available as raw FASTQ files for each sample type (tumor/normal) and SNP array data are available as raw signal intensity files in text format for each sample type (tumor/normal). Access to patient sequence data can be gained for academic use through application to the independent Data Access Committee. Responses to data requests will be provided within two weeks. Information on how to apply for access is available at the EGA under accession code EGAS00001008059. The raw methylation data sets have been submitted to the Gene Expression Omnibus (GEO; https://www.ncbi.nlm.nih.gov/geo/) under accession code GSE292140 (https://www.ncbi.nlm.nih.gov/geo/query/acc.cgi?acc=GSE292140) with no access restrictions. no access restrictions.

### ICGC dataset

Previously published WGS and RNA-seq data generated as part of the ICGC Ovarian Cancer project^58^ are available from the EGA repository as a single bam file for each sample type (tumor/normal), under the accession code EGAD00001000877 (“https://ega-archive.org/datasets/EGAD00001000877“https://ega-archive.org/datasets/EGAD00001000877). Due to the sensitive nature of these patient datasets, access is subject to approval from the ICGC Data Access Compliance Office (https://docs.icgc.org/download/data-access/), an independent body who authorizes controlled access to ICGC sequencing data. ICGC SNP array and methylation data sets have been deposited into the Gene Expression Omnibus (GEO; https://www.ncbi.nlm.nih.gov/geo/) under accession code GSE65821 (https://www.ncbi.nlm.nih.gov/geo/query/acc.cgi?acc=GSE65821), without access restrictions. ICGC gene count level transcriptomic data has been deposited into the GEO under accession code GSE209964 (https://www.ncbi.nlm.nih.gov/geo/query/acc.cgi?acc=GSE209964). ICGC SNP array and methylation data sets have been deposited into GEO (https://www.ncbi.nlm.nih.gov/geo/) under accession code GSE65821 (https://www.ncbi.nlm.nih.gov/geo/query/acc.cgi?acc=GSE65821), without access restrictions. ICGC gene count level transcriptomic data has been deposited into the GEO under accession code GSE209964 (https://www.ncbi.nlm.nih.gov/geo/query/acc.cgi?acc=GSE209964).

### MOCOG dataset

WGS, RNA-seq and SNP array data from long-term survivors generated as part of the MOCOG study^22^ have been deposited in the EGA repository under accession code EGAS00001005984. WGS and RNA-seq data are available as raw FASTQ files for each sample type (tumor/normal) and SNP array data are available as raw signal intensity files in text format for each sample type (tumor/normal). Access to patient sequence data can be gained for academic use through application to the independent Data Access Committee). Responses to data requests will be provided within two weeks. Information on how to apply for access is available at the EGA under accession code EGAS00001005984. The MOCOG cohort raw methylation data sets have been submitted to the GEO under accession code GSE211687 (https://www.ncbi.nlm.nih.gov/geo/query/acc.cgi?acc=GSE211687), with no access restrictions.

### OTTA dataset

Participants of this study did not agree to their data being shared publicly; accordingly, the data used in this research will not be made available.

Uniformly processed somatic variant data from the ICGC, MOCOG, and short survival *BRCA* cohorts is deposited in Synapse under accession code syn65463502 and processed expression and methylation data from all cohorts has been submitted into the GEO under accession code GSE292140 (https://www.ncbi.nlm.nih.gov/geo/query/acc.cgi?acc=GSE292140) and GSE292142 (https://www.ncbi.nlm.nih.gov/geo/query/acc.cgi?acc=GSE292142), without access restrictions. All other data are available within the article (and its Supplementary Information files) or from the corresponding authors on request.

Population frequencies of genetic variants can be accessed via the Genome Aggregation Database (gnomAD) at https://gnomad.broadinstitute.org/. Supporting evidence for pathogenicity of genomic alterations can be accessed via ClinVar (https://www.ncbi.nlm.nih.gov/clinvar/), BRCA Exchange (https://brcaexchange.org/) and the *TP53* Database (https://tp53.isb-cgc.org/). The Ensembl ranked order of severity of variant consequences is available at: https://m.ensembl.org/info/genome/variation/prediction/predicted_data.html. Mutational signature reference databases can be accessed via COSMIC (https://cancer.sanger.ac.uk/signatures/) and Signal (https://signal.mutationalsignatures.com/). The LM22 signature matrix used for immune cell deconvolution can be downloaded here: https://cibersortx.stanford.edu/. MSigDB hallmark gene sets can be accessed here: https://www.gsea-msigdb.org/gsea/msigdb/. Illumina methylation probes that were filtered out due to poor performance (e.g. cross reactive or non-specific probes) can be found here: https://github.com/sirselim/illumina450k_filtering. Germline polymorphic sites for reference and variant allele read counts used in FACETS analysis can be found at ftp://ftp.ncbi.nih.gov/snp/organisms/human_9606_b151_GRCh37p13/VCF/common_all_20180423.vcf.gz. The GTF used for annotation and RNA-seq counts is available here: ftp://ftp.ensembl.org/pub/grch37/release-92/.

## Code availability

No custom code or software was used in the data analyses and for the figures. All results can be replicated using publicly available tools and software. The tools and versions used are described in the Methods and Supplementary Information.

## Acknowledgments

We thank A. Freimund, R. Lupat, J. Ellul, and the Peter MacCallum Cancer Centre Research Computing Facility for their contributions to the study. This work was supported by the National Health and Medical Research Council (NHMRC) of Australia (GNT1186505 and GNT2029088), the US Army Medical Research and Materiel Command Ovarian Cancer Research Program (Award No. W81XWH-16-2-0010 and W81XWH-21-1-0401), the National Institutes of Health (NIH) (R21-CA267050, K07-CA080668, R01-CA95023, R01-CA248288, P50-CA136393, P30-CA015083, MO1-RR000056), the Swiss National Foundation (P500PM_20726); Bangerter-Rhyner Stiftung (0297); Margarete and Walter Lichtenstein-Stiftung; and Freie Gesellschaft Basel. The Gynaecological Oncology Biobank at Westmead was funded by the NHMRC (ID310670, ID628903); the Cancer Institute NSW (12/RIG/1-17, 15/RIG/1-16); the Department of Gynaecological Oncology, Westmead Hospital; and acknowledges financial support from the Sydney West Translational Cancer Research Centre, funded by the Cancer Institute NSW (15/TRC/1-01). Direct funding for the generation of the NanoString data for OTTA was provided by the NIH (R01-CA172404, and R01-CA168758), the Canadian Institutes for Health Research (Proof-of-Principle I program) and the United States Department of Defense Ovarian Cancer Research Program (OC110433). T.A.Z. is supported by the Swiss National Foundation Return CH Postdoc.Mobility (P5R5PM_222151). D.W.G. is supported by a Victorian Cancer Agency/Ovarian Cancer Australia Low-Survival Cancer Philanthropic Mid-Career Research Fellowship (MCRF22018) and the Ovarian Cancer Research Foundation (2025/OCRF0071). S.J.R. is supported by the NHMRC (2009840). M.J.G is supported by the Ministerio de Ciencia, Innovación y Universidades (MICIU)/AEI/10.13039/501100011033 and ERDF, EU (Project PID2023-151298OB-I00). A.O. is partially funded by Ministerio de Ciencia e Innovación, Instituto de Salud Carlos III (PI23/01235) supported by FEDER funds and the Spanish Network on Rare Diseases (CIBERER). K.M.D., T.P.C., and G.L.M. were supported by awards from the Uniformed Services University of the Health Sciences and the Defense Health Program to the Henry M Jackson Foundation (HJF) for the Advancement of Military Medicine Inc. to the Gynecologic Cancer Center of Excellence Program including HU0001-16-2-0006 (PIs: Chad A. Hamilton and G. Larry Maxwell), HU0001-19-2-0031, HU0001-20-2-0033, and HU0001-21-2-0027 (PIs: Yovanni Casablanca and G. Larry Maxwell), HU0001-22-2-0016 and HU0001-23-2-0038 (PIs: Neil T. Phippen and G. Larry Maxwell), as well as HU0001-23-2-0038 and HU0001-24-2-0047 (PIs Christopher M Tarney and G. Larry Maxwell). T.V.G. is a Senior Clinical Investigator of the Fund for Scientific Research-Flanders (FWO Vlaanderen 18B2921N). A.DeF. is supported by the NHMRC (2033042). The AOV study was funded by the Canadian Institutes for Health Research (MOP-86727). The Generations Study was funded by Breast Cancer Now and the United Kingdom National Health Service funding to the Royal Marsden/Institute of Cancer Research. The UK Ovarian Cancer Population study (UKOPS) was funded by The Eve Appeal (The Oak Foundation) with contribution to authors’ salary through MRC core funding MC_UU_00004/01 and the NIH Research University College London Hospitals Biomedical Research Centre. The contents of the published material are solely the responsibility of the authors and do not reflect the views of the NHMRC, NIH, and other funders.

## Authors contributions

T.A.Z.: Conceptualization, data curation, formal analysis, funding acquisition, validation, investigation, visualization, methodology, writing–original draft, writing–review and editing. S.F.: Conceptualization, data curation, validation, methodology, writing–original draft, writing–review and editing. A.P.: Conceptualization, data curation, formal analysis, validation, investigation, visualization, methodology, writing–original draft, writing–review and editing. D.A.: Data curation, validation, investigation, visualization, methodology, writing–original draft, writing–review and editing. M.W.J.: Formal analysis, validation, investigation, visualization, methodology, writing–original draft, writing–review and editing. L.T.: Formal analysis, validation, investigation, visualization, methodology, writing–original draft, writing– review and editing. A.F.: Conceptualization, data curation, investigation, writing–review and editing. C.M.L.: Formal analysis, validation, investigation, methodology, writing–review and editing. C.J.K.: Resources, data curation, methodology, writing–original draft, writing–review and editing. A.B.: Resources, writing–review and editing. N.S.M.: Resources, writing–review and editing. K.M.: Resources, writing–review and editing. P.H.: Data curation, formal analysis, validation, investigation, methodology, writing–review and editing. J.A.: Resources, writing– review and editing. A.C.A.: Resources, writing–review and editing. G.A-Y.: Resources, writing–review and editing. M.W.B.: Resources, writing–review and editing. A.B.: Resources, writing–review and editing. C.B.: Resources, writing–review and editing. F.B.: Resources, writing–review and editing. C.B.: Resources, writing–review and editing. J.B.: Resources, writing–review and editing. A.H.B.: Resources, writing–review and editing. M.E.C.: Resources, writing–review and editing. A.C-J.: Resources, writing–review and editing. D.S.C.: Resources, writing–review and editing. E.L.C.: Resources, writing–review and editing. A.C-G.: Resources, writing–review and editing. P.C.: Resources, writing–review and editing. K.L.C-H.: Resources, writing–review and editing. C.C.: Resources, writing–review and editing. K.M.D.: Resources, writing–review and editing. C.D.: Resources, writing–review and editing. T.D.: Resources, writing–review and editing. A.B.E.: Resources, writing–review and editing. E.E.: Resources, writing–review and editing. J.E.: Resources, writing–review and editing. T.E.: Resources, writing–review and editing. R.F.: Resources, writing–review and editing. A.F.: Resources, writing–review and editing. M.G-C.: Resources, writing–review and editing. A.G-M.: Resources, writing–review and editing. P.G.: Resources, writing–review and editing. R.G.: Resources, writing–review and editing. P.H.: Resources, writing–review and editing. A.D.H.: Resources, writing–review and editing. A.H.: Resources, writing–review and editing. S.H.: Resources, writing–review and editing. B.Y.H.: Resources, writing–review and editing. A.H.: Resources, writing–review and editing. S.H.: Resources, writing–review and editing. D.G.H.: Resources, writing–review and editing. M.J-L.: Resources, writing–review and editing. M.E.J.: Resources, writing–review and editing. E.K.: Resources, writing–review and editing. E.K.: Resources, writing–review and editing. T.K.: Resources, writing–review and editing. F.K.F.K.: Resources, writing–review and editing. G.K.: Resources, writing–review and editing. R.F.P.M.K.: Resources, writing–review and editing. J.K.: Resources, writing– review and editing. D.L.: Resources, writing–review and editing. C-H.L.: Resources, writing– review and editing. J.L.: Resources, writing–review and editing. S.C.Y.L.: Resources, writing– review and editing. Y.L.: Resources, writing–review and editing. A.L.: Resources, writing– review and editing. J.L.: Resources, writing–review and editing. L.L.: Resources, writing– review and editing. J.L.: Resources, writing–review and editing. C.M.: Resources, writing– review and editing. I.A.M.: Resources, writing–review and editing. M.M.: Resources, writing– review and editing. G.S.N.: Resources, writing–review and editing. N.N.: Resources, writing– review and editing. A.O.: Resources, writing–review and editing. S.O.: Resources, writing– review and editing. A.O.: Resources, writing–review and editing. C.M.Q.: Resources, writing– review and editing. G.RM.: Resources, writing–review and editing. I.R-C.: Resources, writing– review and editing. C.R-A.: Resources, writing–review and editing. P.R.: Resources, writing– review and editing. M.R.: Resources, writing–review and editing. S.G.S.: Resources, writing– review and editing. S.S.: Resources, writing–review and editing. M.J.S.: Resources, writing– review and editing. H-P.S.: Resources, writing–review and editing. G.S.S.: Resources, writing– review and editing. L.S.: Resources, writing–review and editing. C.J.R.S.: Resources, writing– review and editing. A.T.: Resources, writing–review and editing. A.T.: Resources, writing– review and editing. C.M.T.: Resources, writing–review and editing. S.E.T.: Resources, writing–review and editing. K.K.V.: Resources, writing–review and editing. M.A.A.: Resources, writing–review and editing. T.G.: Resources, writing–review and editing. E.N.: Resources, writing–review and editing. L.W.: Resources, writing–review and editing. A.E.W-H.: Resources, writing–review and editing. C.W.: Resources, writing–review and editing. C.W.: Resources, writing–review and editing. J.W.: Resources, writing–review and editing. N.W.: Resources, writing–review and editing. L.R.W.: Resources, writing–review and editing. S.J.W.: Resources, writing–review and editing. B.W.: Resources, writing–review and editing. M.S.A.: Resources, writing–review and editing. A.B.: Resources, writing–review and editing. F.J.C-R.: Resources, writing–review and editing. P.A.C.: Resources, writing–review and editing. T.P.C.: Resources, writing–review and editing. P.C.: Resources, writing–review and editing. J.A.D.: Resources, writing–review and editing. P.A.F.: Resources, writing–review and editing. R.T.F.: Resources, writing–review and editing. M.J.G.: Resources, writing–review and editing. S.A.G.: Resources, writing–review and editing. M.T.G.: Resources, writing–review and editing. J.G.: Resources, writing–review and editing. H.R.H.: Resources, writing–review and editing. F.H.: Resources, writing–review and editing. H.M.H.: Resources, writing–review and editing. B.Y.K.: Resources, writing–review and editing. L.E.K.: Resources, writing– review and editing. G.L.M.: Resources, writing–review and editing. U.M.: Resources, writing– review and editing. F.M.: Resources, writing–review and editing. S.L.N.: Resources, writing– review and editing. J.M.S.: Resources, writing–review and editing. A.S.: Resources, writing– review and editing. A.J.S.: Resources, writing–review and editing. I.V.: Resources, writing– review and editing. A.H.W.: Resources, writing–review and editing. J.D.B.: Resources, writing–review and editing. P.D.P.P.: Resources, writing–review and editing. C.L.P.: Resources, writing–review and editing. M.C.P.: Resources, writing–review and editing. E.L.G.: Resources, writing–review and editing. S.J.R.: Conceptualization, resources, data curation, supervision, funding acquisition, validation, writing–original draft, project administration, writing–review and editing. M.K.: Conceptualization, resources, data curation, investigation, formal analysis, validation, visualization, supervision, methodology, writing– original draft, writing–review and editing. B.N.: Resources, data curation, investigation, formal analysis, validation, visualization, methodology, writing–review and editing. A.DF.: Conceptualization, Resources, writing–review and editing. M.L.F.: Conceptualization, Resources, writing–review and editing. D.D.L.B.: Conceptualization, resources, supervision, funding acquisition, writing–original draft, writing–review and editing. D.W.G.: Conceptualization, resources, data curation, formal analysis, supervision, funding acquisition, validation, investigation, visualization, methodology, writing–original draft, project administration, writing–review and editing.

## Extended Data Figure legends

**Extended Data Fig. 1 | Patient cohorts and case selection: Overview for clinical, molecular, and validation analysis.** Overview of patient cohorts and case selection for the clinical, molecular and validation analysis.

*HGSC=Tubo-ovarian high-grade serous carcinoma, AOCS =Australian Ovarian Cancer Study, MOCOG=Multidisciplinary Ovarian Cancer Outcome Group, gBRCApv=pathogenic germline BRCA variant, mcIF=multicolor immunofluorescence, OTTA = Ovarian Tumor Tissue Analysis, mRNA=messenger ribonucleic acid, OS=overall survival*

**Extended Data Fig. 2 | Association of *BRCA* status and residual disease with survival and distribution of molecular features in HGSC: Insights from the OTTA cohort. a,** Multivariable Cox proportional hazards model of the interaction term *BRCA* and residual disease status and clinicopathological and molecular predictive features on overall survival with patients from the OTTA cohort. *P* values were derived using the Wald test; values < 0.05 are colored red (*, *P* < 0.05; **, *P* < 0.01; ***, *P* < 0.001; \*\*\*\**P* < 0.0001). **b**, Distribution of molecular features (CD8+ TIL density, molecular subtypes, and *RB1* loss) within the *BRCA* and residual groups by odds ratios. *P*-values were calculated based on odds ratios (OR).

*R=Residual disease, R0=No residual disease, gBRCApv=pathogenic germline BRCA variant, HR=Hazard ratio, CI=confidence interval, TIL= tumor-infiltrating lymphocyte*

**Extended Data Fig. 3 | Association of *BRCA* status and neoadjuvant chemotherapy on survival in HGSC.** Kaplan-Meier analysis of overall survival stratified by the interaction term *BRCA* and neoadjuvant chemotherapy status from patients of the Australian Ovarian Cancer Study (AOCS) cohort. *P* value calculated by log-rank test.

*gBRCApv=pathogenic germline BRCA variant, NACT=neoadjuvant chemotherapy, n=Number of patients, OS=Overall survival*

**Extended Data Fig. 4 | Prognostic significance of pathogenic germline *BRCA1* and *BRCA2* variant by domain location and mutation type on outcome in HGSC. a,** and **b**, shows the distribution of pathogenic germline mutations on the *BRCA1* and *BRCA2* gene, respectively. **c**, and **d**, show the distribution of mutation types within the different *BRCA1* and *BRCA2* functional domains. Fisher’s exact test *P* value is reported. *gBRCApv=pathogenic germline*

*BRCA variant, DBD=DNA binding domain, RING=Really Interesting New Gene domain, RAD51-BD=RAD51-binding domain, BRCT=BRCA c-terminal domain, n=Number of patients*

**Extended Data Fig. 5 | Distribution analysis of clinical and molecular features by *BRCA* and survival groups in HGSC. a**, Box plots summarizing numerical, clinical and genomic characteristics by *BRCA* and survival groups; dots represent each sample, boxes show the interquartile range (25-75th percentiles), central lines show the median, whiskers show the smallest/largest values within 1.5 times the interquartile range. Kruskal-Wallis test Benjamini-Hochberg adjusted *P*-values and pairwise Mann-Whitney-Wilcoxon test *P* values (two sided) are reported (ns, *P*>0.1; ., *P*<0.1; *, *P* < 0.05; **, *P* < 0.01; ***, *P* < 0.001). Features are ranked according to their significance. **b**, Proportion of patients with categorical characteristics per *BRCA* and survival group. Features are ordered by significance using Fisher’s exact test (two-tailed) and clusters are ordered by proportion of long-term survivors. Fisher’s test *P*-values shown are Benjamini-Hochberg adjusted *P*-values. Features are ordered by significance.

*LST=Large scale transitions, LOH= Loss of heterozygosity, SV= Structural variants, DEL=Deletion, DUP=Duplication, AI=Allelic imbalance, INV=Inversion; Long term survivor (LTS)= OS >3 years, Short term survivor (STS)= OS ≤3 years, BRCA-P=BRCA-proficient; HR status= Homologous recombination status, P=Progression, PF= Progression-free; R=residual disease, R0=no residual disease; High ≥63, Moderate= 42-62, Low= ≤41, IMMB=Immune clusters (by CIBERSORTx), Molecular subtypes: C1.MES=C1 mesenchymal subtype, C2.IMM=C2 immunoreactive subtype, C4.DIF=C4 differentiated subtype, C5.PRO=C5 proliferative subtype*

**Extended Data Fig. 6 | *NF1* gene alterations and expression. a,** Scatter graphs (right) show *NF1* expression (y-axis) plotted against copy number (x-axis) in primary tumors (n=153, Spearman correlation analysis). Boxplots (left) summarize *NF1* expression by *NF1* alterations with and without locus specific loss of heterozygosity (LOH); lines indicate median, and whiskers show range. Kruskal–Wallis test *P* value is reported as well as pairwise Wilcoxon rank-sum test *P* values comparing altered groups to wildtype (non-significant (ns), *P*>0.05; ****, *P* <0.0001). **b**, Kaplan-Meier analysis of overall survival in patients stratified by *NF1* alterations exhibiting locus specific LOH and **c**, in patients with *BRCA1*-and *BRCA2*-deficient tumors stratified by *NF1* alteration status. *P* values calculated by log-rank test.

*TMM=Trimmed Mean of M-values*

**Extended Data Fig. 7 | Survival analysis by *NF1* expression HGSC, with stratification by *BRCA* status: Findings from MOCOG and OTTA cohorts. Kaplan-Meier curves for overall survival (OS) comparing a** patients with HGSC from the MOCOG cohort by NF1 protein expression status (NF1 retained vs loss) and in **b** additionally stratified by germline *BRCA* mutation status. **c**, Kaplan-Meier curve comparing the overall survival of patients with HGSC from the OTTA cohort by *NF1* RNA expression status (low=lowest quantile, high=2^nd^ to 5^th^ quantiles) and in **d** additionally stratified by germline *BRCA* mutation status. *P* values calculated by log-rank test

**Extended Data Fig. 8 | *PIK3CA* and *RAD21* gene alterations in HGSC. a, b,** Scatter graphs (right) of the expression (y-axis) of *PIK3CA* (a) and *RAD21* (b) plotted against copy number (x-axis) in primary tumors (n=153, Spearman correlation analysis). Boxplots (left) summarize expression by mutation type; lines indicate median, and whiskers show range. Kruskal–Wallis test *P* value is reported as well as pairwise Wilcoxon rank-sum test *P* values comparing altered groups to wildtype (non-significant (ns), *P*>0.05; ****, *P* <0.0001; ***, *P* <0.001; **, *P* <0.01). **c**, Kaplan-Meier analysis of overall survival in patients with HGSC stratified by *BRCA*-status and *PIK3CA* amplification vs no amplification*. P* value calculated by log-rank test. **d**, Kaplan-Meier analysis of overall survival in patients with HGSC stratified by *BRCA*-status and *RAD21* amplification vs no amplification. *P* value calculated by log-rank test. **e**, Kaplan-Meier analysis of overall survival in patients with HGSC from the OTTA cohort stratified by *PIK3CA* RNA expression status (high=highest quantile, low=1^st^ to 4^th^ quantiles) and stratified by germline *BRCA* mutation status. *P* value calculated by log-rank test.

*SV=Structural variants, amp=amplification, WG=Whole gene, BRCA-P=BRCA-proficient, TMM=Trimmed Mean of M-values*

**Extended Data Fig. 9 | *c-KIT* gene expression in HGSC: Association with survival, molecular subtypes, and *BRCA* status. a**, Forest plot (left) indicates the hazard ratio (HR, squares) and 95% confidence interval (CI; whiskers) for overall survival (OS) calculated using a multivariable Cox proportional hazard regression model based on the LM22 immune cell types detected by CIBERSORTx analysis (n = 153 patients). Cell types are arranged by HR. *P* values were derived by Wald test; values < 0.05 are colored red (\**P* < 0.05, \*\**P* < 0.01). **b**, Differential expression analysis was performed using DESeq2 to determine fold change (right) of gene expression between the *BRCA* survival groups (*BRCA1=BRCA1*-deficient; *BRCA2=BRCA2*-deficient; *BRCA-P*=*BRCA*-proficient; Long term survivor (LTS) = OS >3 years; Short term survivor (STS) = OS ≤3 years) (two-tailed Wald test, both unadjusted *P* values and Benjamini-Hochberg adjusted *P* values (*P*_adj_) are shown). **c**, Multivariable Cox proportional hazards model for OS comparing *c-KIT* with high vs low RNA expression levels by median and adjusted for HRD sum score and BRCA HRD status. *P* values were derived by Wald test; values < 0.05 are colored red (\**P* < 0.05, \*\**P* < 0.01, \*\*\**P* < 0.001, \*\*\*\**P* < 0.0001). **d**, Boxplots summarize RNA expression of the *c-KIT* gene marker across the molecular subtypes (C1.MES=C1 mesenchymal subtype, C2.IMM=C2 immunoreactive subtype, C4.DIF=C4 differentiated subtype, C5.PRO=C5 proliferative subtype); points represent each sample, boxes show the interquartile range (25–75th percentiles), central lines indicate the median, and whiskers show the smallest/largest values within 1.5 times the interquartile range. Differential expression analysis was performed using DESeq2 to determine fold change (right) of gene expression between the molecular subtypes (two-tailed Wald test, both unadjusted *P* values and Benjamini-Hochberg adjusted *P* values (*P*_adj_) are shown). **e**, Clustered heatmap summarizing gene set enrichment analysis (GSEA) using the hallmark Molecular Signatures Database (MSigDB) gene sets. Direction and color of triangles relate to the normalized enrichment score (NES) as generated by FGSEA. *P* values (two-sided) were calculated using the FGSEA default Monte Carlo method; the size of the triangles corresponds to the negative log_10_ Benjamini-Hochberg adjusted *P* value (*P*_adj_). Columns are separated by molecular subtypes with the direction of enrichment indicated by the first group mentioned in the x-axis label.

*BRCA-P= BRCA-proficient, Survival group: Long-term survivor (LTS)= OS >3 years, Short-term survivor (STS)= OS ≤3 years, TMM=Trimmed Mean of M-values*

## Supplementary Tables

**Table S1** Clinical data of the AOCS cohort

**Table S2** Mutation type and location g*BRCA*pv-carriers AOCS cohort

**Table S3** Univariable and multivariable Accelerated Failure Time (AFT) model results of clinical features, including interaction analyses with gBRCApv status, on overall survival in patients with HGSC from the AOCS cohort.

**Table S4** Univariable and multivariable Accelerated Failure Time (AFT) model results of clinical features, including interaction analyses with gBRCApv status, on progression-free survival in patients with HGSC from the AOCS cohort.

**Table S5** Distribution of molecular subtypes, tumor-infiltrating lymphocytes, RB1 protein expression status, and *CCNE1* amplification status stratified by residual disease or no residual disease, and by g*BRCA*pv carrier or non-carrier status. Percentages are given in parentheses and calculated within each subgroup.

**Table S6** Univariable and multivariable Accelerated Failure Time (AFT) model of gBRCApv and neoadjuvant chemotherapy status and clinicopathological predictive features on overall survival in patients with HGSC from the AOCS cohort.

**Table S7** Clinical data of the multi-omics cohort

**Table S8** Mutation location with Δ11q proportion and Δ11q *BRCA1* expression

**Table S9** Results of the Kaplan–Meier analysis of overall survival in 154 patients with HGSC from the multi-omics cohort stratified by the main features of interest.

**Table S10** *BRCA* groups, HRD score, CHORD scores, whole genome duplication, and molecular signatures multi-omics cohort

**Table S11** Germline alterations in genes of interest with loss of wildtype allele and clonality

**Table S12** Somatic alterations in genes of interest

**Table S13** Frequency of genes identified as significant in both differential methylation and differential expression analyses.

**Table S14** *NF1* alteration type, segment copy number, clonality, loss of heterozygosity and, and RNA expression.

**Table S15** Mutual exclusivity and co-occurrence analysis whole multi-omics cohort

**Table S16** Mutual exclusivity and co-occurrence analysis short survival *BRCA* group

**Table S17** *PIK3CA* segment copy number, RNA expression, and alteration type

**Table S18** *RAD21* segment copy number, RNA expression, and alteration type

**Table S19** Mutation and neoantigen burden

**Table S20** Quartile odds ratios of immune cell subsets comparing short-term (n=36) vs long-term (n=106) survival group

**Table S21** Relative CIBERSORTx abundance of LM22 cell types of the multi-omics cohort with cluster

**Table S22** Details of participating study sites and ethics approvals from the AOCS, MOCOG and OTTA cohort.

