## Extended Data Figures for "Beyond *BRCA* deficiency: Clinical and molecular predictors of survival in patients with *BRCA*-deficient tubo-ovarian high-grade serous carcinoma"

**Extended Data Figure 1**

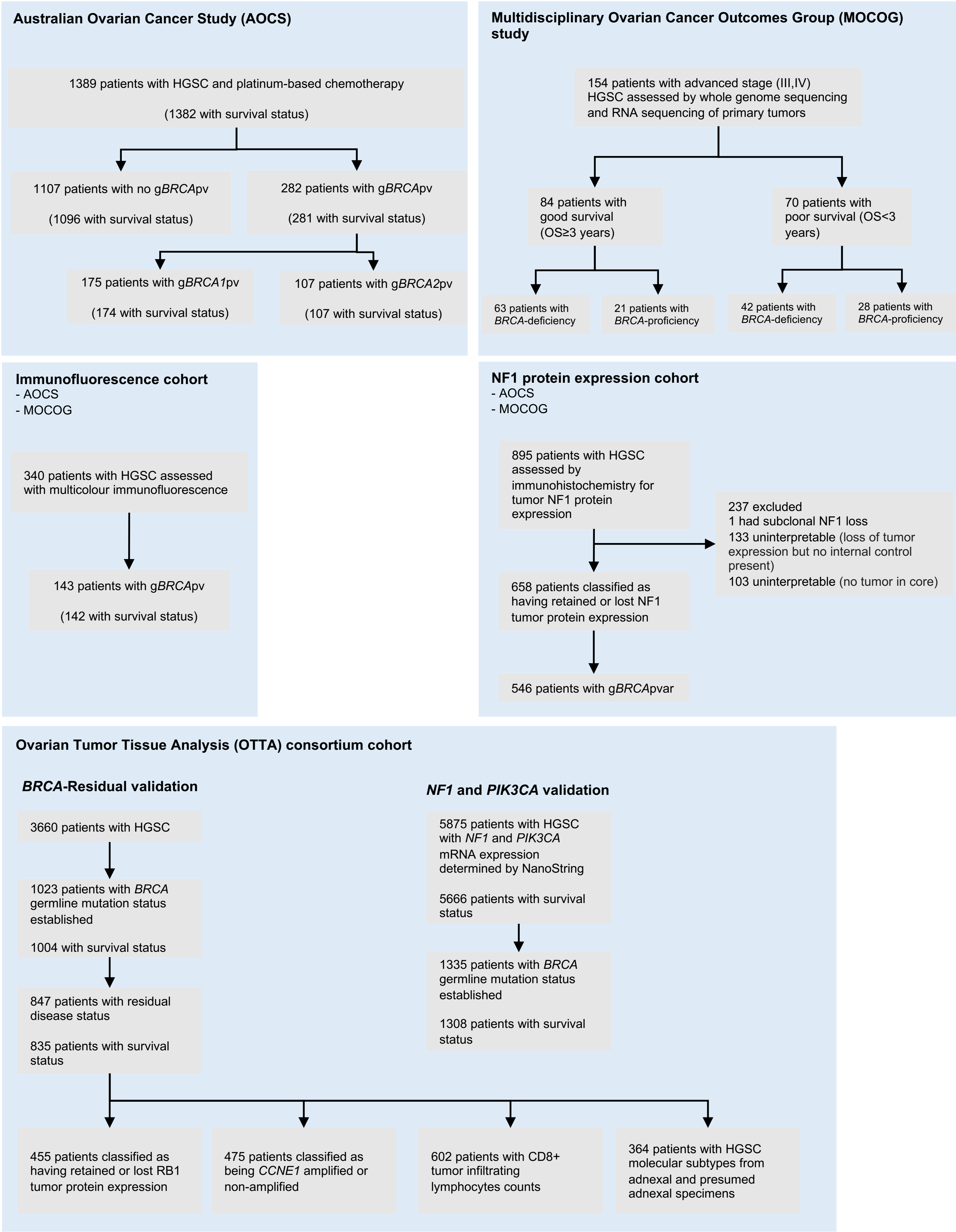

**Extended Data Figure 2**

**A.**

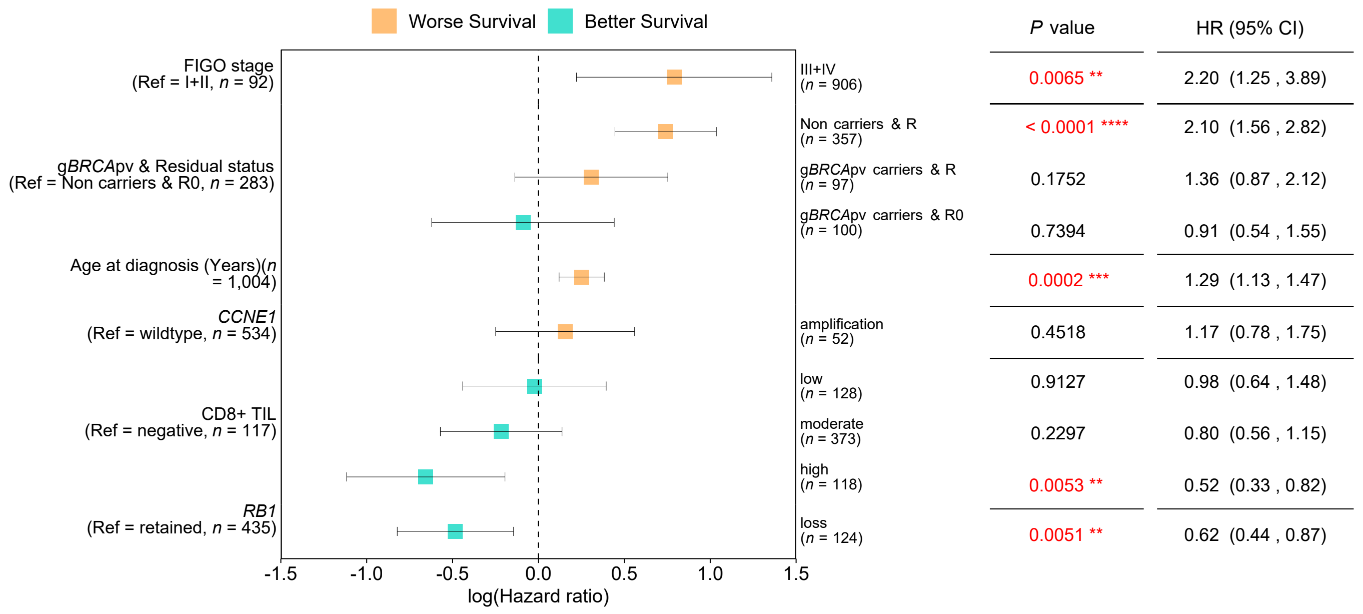


**B.**

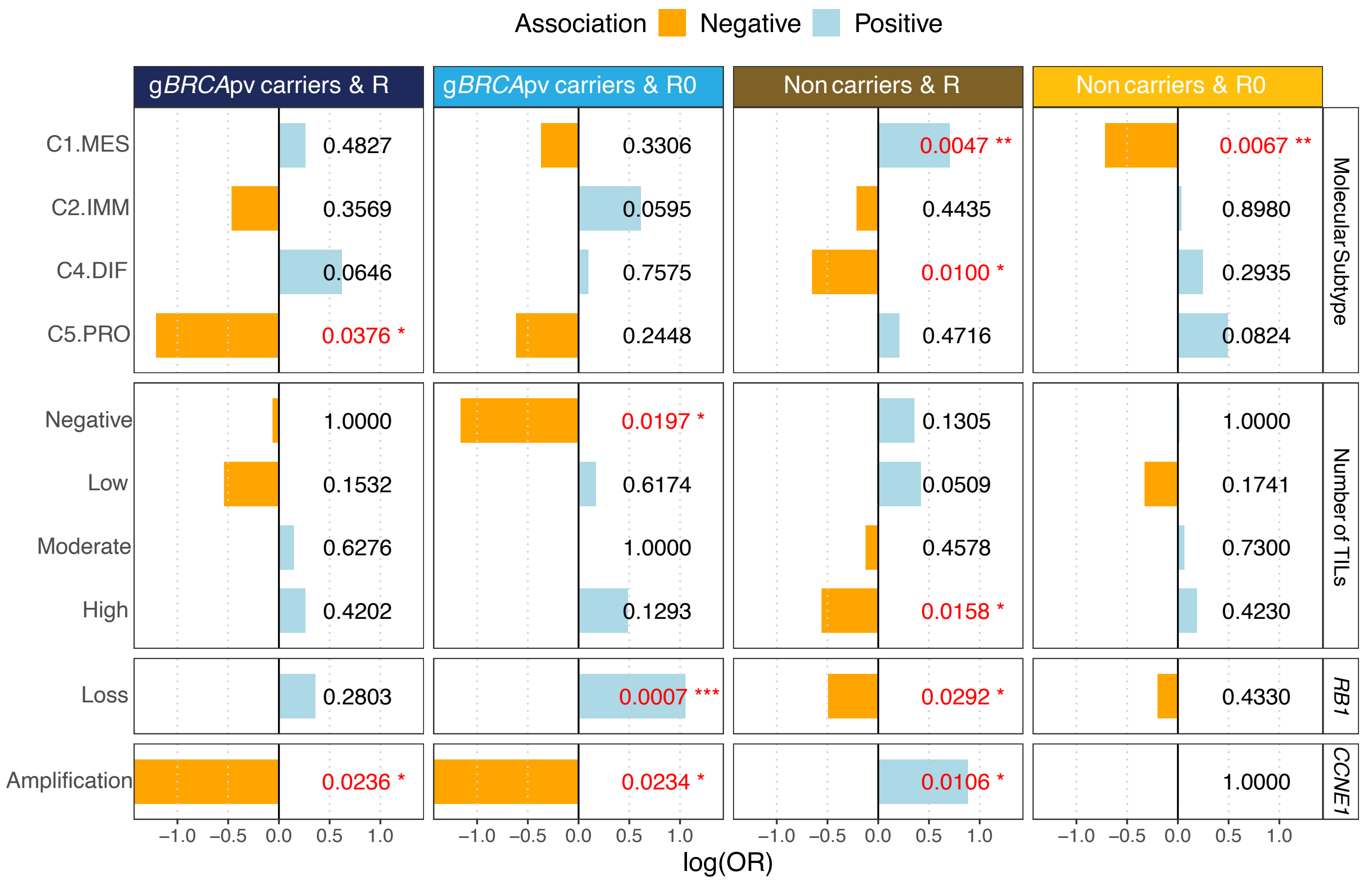

**Extended Data Figure 3**

| | *gBRCA*pv & no NACT (n=220) | *gBRCA*pv & NACT (n=54) | Non-carriers & no NACT (n=828) | Non-carriers & NACT (n=268) |
|---|---|---|---|---|
| **Events**, n (%) | 123 (55.9) | 31 (57.1) | 598 (72.2) | 170 (63.4) |
| **Median OS**, years | 6.21 | 4.46 | 3.80 | 3.10 |
| **5-Year OS rate**, % | 57.8 | 43.1 | 40.5 | 30.9 |

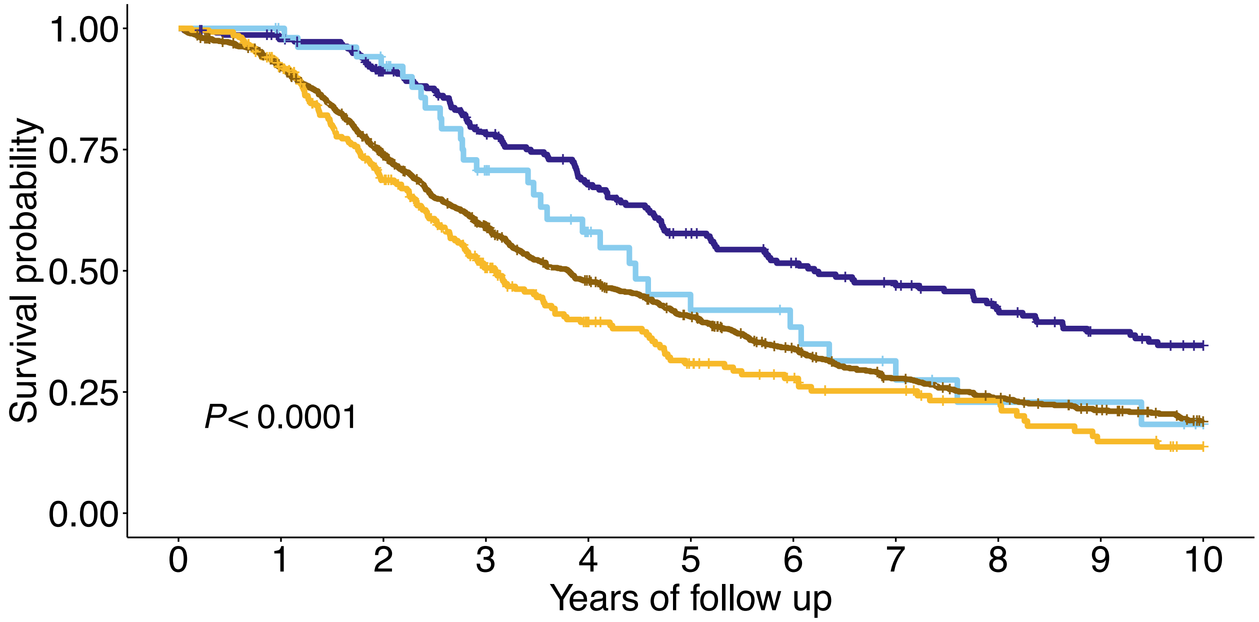


**Number at risk**

| | 0 | 1 | 2 | 3 | 4 | 5 | 6 | 7 | 8 | 9 | 10 |
|---|---|---|---|---|---|---|---|---|---|---|---|
| *gBRCA*pv & no NACT | 220 | 210 | 185 | 154 | 130 | 106 | 91 | 79 | 66 | 54 | 46 |
| *gBRCA*pv & NACT | 54 | 52 | 46 | 31 | 19 | 13 | 11 | 7 | 5 | 5 | 2 |
| Non-carriers & no NACT | 828 | 741 | 579 | 449 | 354 | 288 | 222 | 174 | 145 | 120 | 93 |
| Non-carriers & NACT | 268 | 237 | 165 | 100 | 63 | 45 | 34 | 28 | 22 | 14 | 9 |

**Extended Data Figure 4**

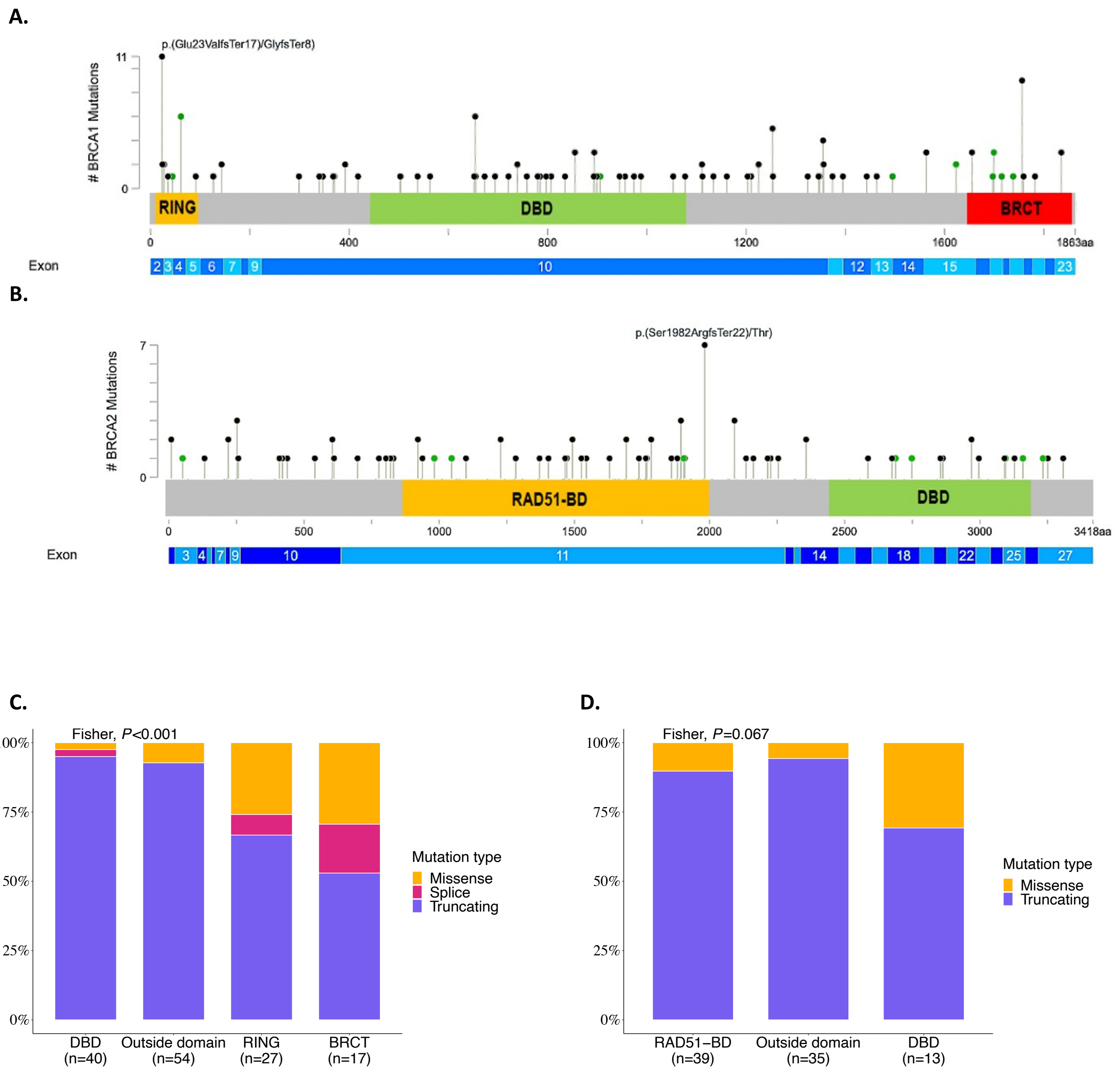

#### Extended Data Figure 5

**A.**

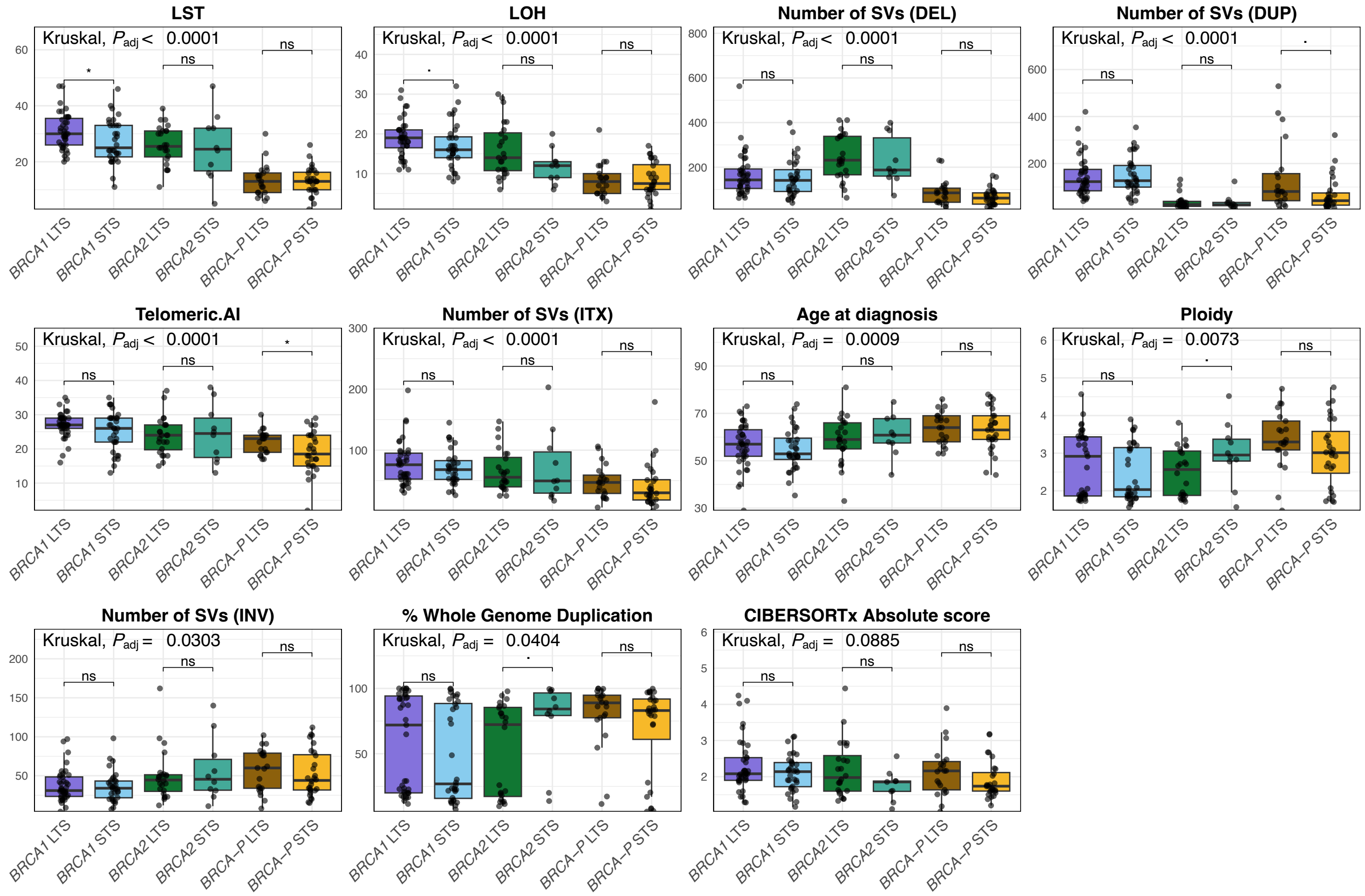


**B.**

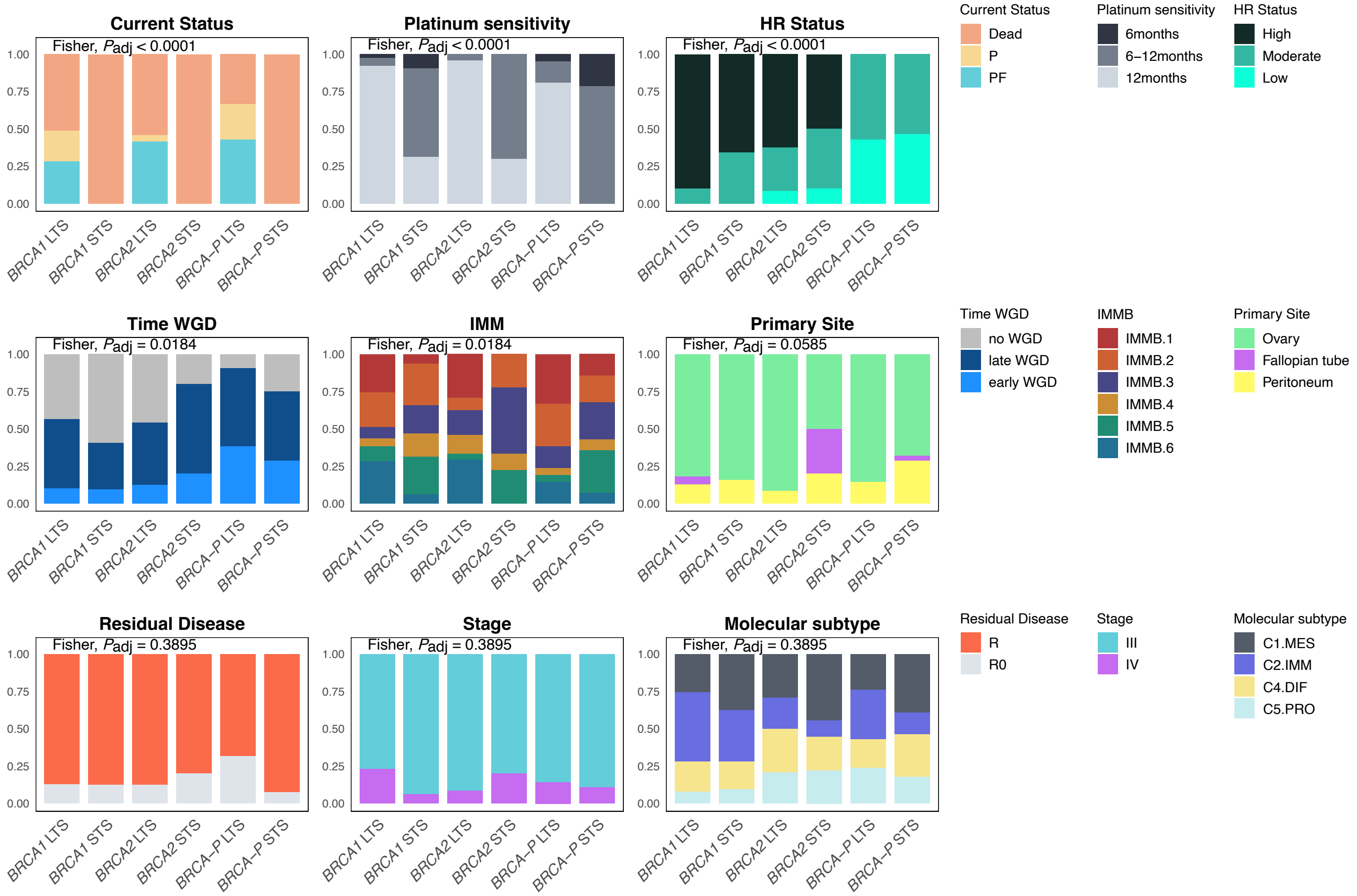

**Extended Data Figure 6**

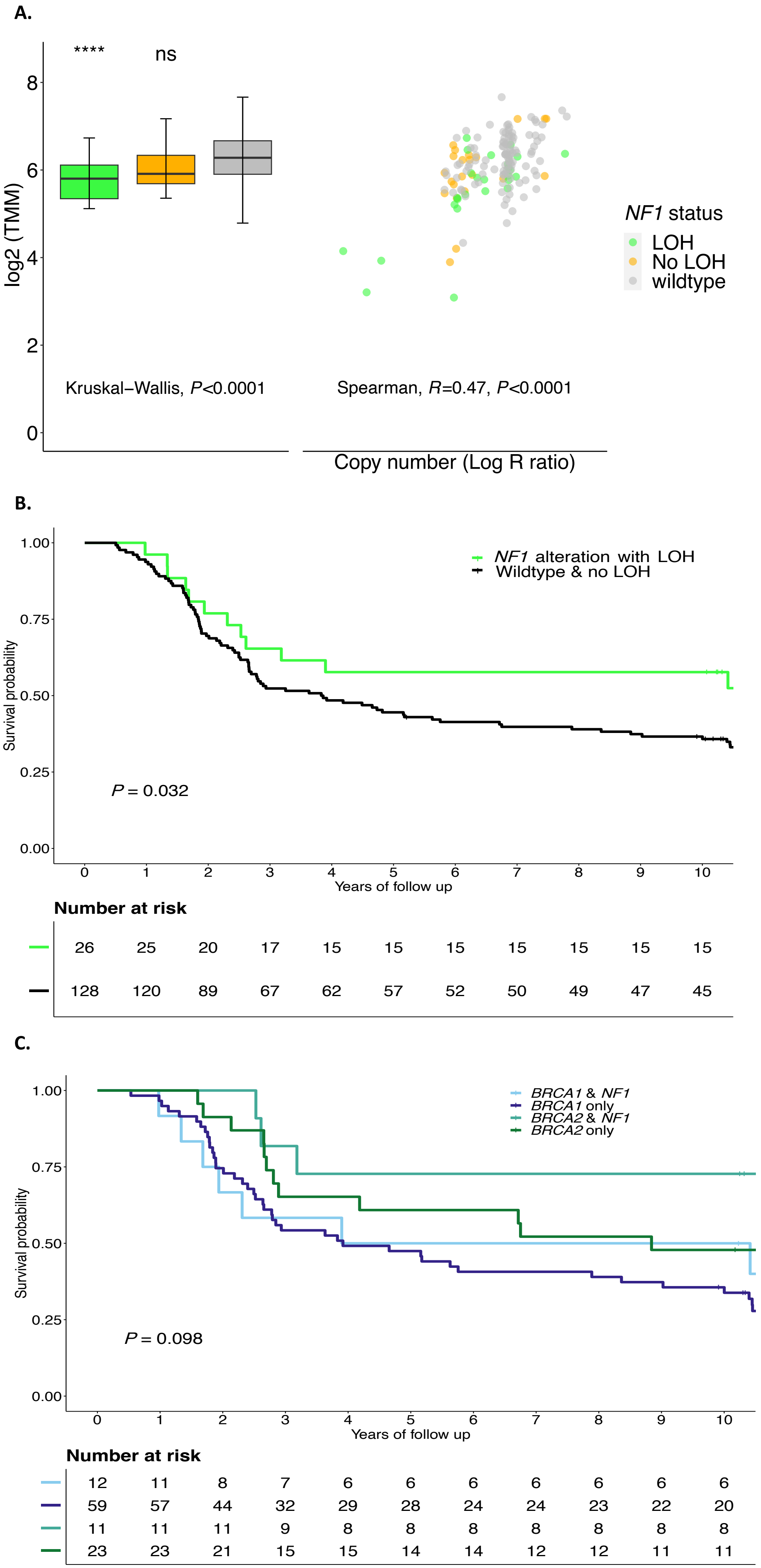

### Extended Data Figure 7

**A.**

| Protein expression | NF1 retained (n=570) | NF1 loss (n=88) |
|---|---|---|
| **Events**, n (%) | 471 (82.6) | 64 (72.7) |
| **Median OS**, years | 3.58 | 4.70 |
| **5-Year OS rate**, % | 35.8 | 47.7 |

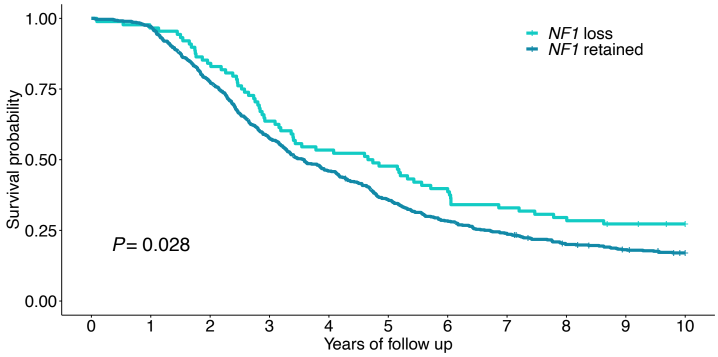


**Number at risk**

| | 0 | 1 | 2 | 3 | 4 | 5 | 6 | 7 | 8 | 9 | 10 |
|---|---|---|---|---|---|---|---|---|---|---|---|
| | 88 | 85 | 74 | 56 | 47 | 42 | 35 | 29 | 26 | 23 | 21 |
| | 570 | 554 | 442 | 328 | 262 | 203 | 161 | 134 | 110 | 97 | 87 |

**B.**

| | *BRCA1* & NF1 retained (n=79) | *BRCA1* & NF1 loss (n=21) | *BRCA2* & NF1 retained (n=50) | *BRCA2* & NF1 loss (n=6) | *BRCA*wt & NF1 retained (n=336) | *BRCA*wt & NF1 loss (n=54) |
|---|---|---|---|---|---|---|
| **Events**, n (%) | 58 (73.4) | 15 (71.4) | 34 (68) | 4 (66.7) | 284 (84.5) | 38 (70.4) |
| **Median OS**, years | 4.69 | 4.74 | 5.72 | 8.05 | 3.36 | 5.01 |
| **5-Year OS rate**, % | 46.8 | 47.6 | 55.9 | 66.7 | 33.9 | 50 |

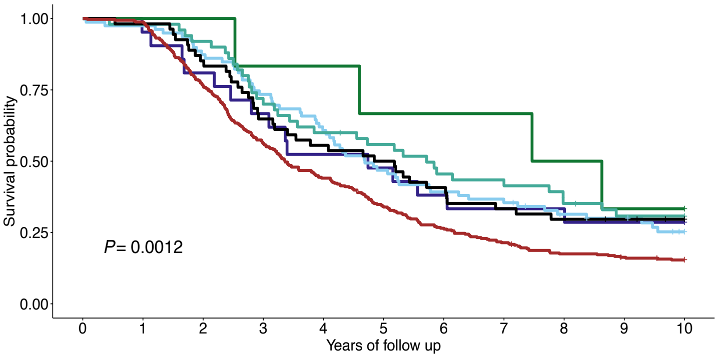


**Number at risk**

| | 0 | 1 | 2 | 3 | 4 | 5 | 6 | 7 | 8 | 9 | 10 |
|---|---|---|---|---|---|---|---|---|---|---|---|
| | 21 | 20 | 17 | 14 | 11 | 10 | 8 | 7 | 7 | 6 | 6 |
| | 79 | 77 | 69 | 58 | 48 | 37 | 31 | 28 | 22 | 20 | 14 |
| | 6 | 6 | 6 | 5 | 5 | 4 | 4 | 4 | 3 | 2 | 2 |
| | 50 | 49 | 46 | 35 | 30 | 27 | 22 | 20 | 17 | 14 | 14 |
| | 54 | 53 | 46 | 35 | 30 | 27 | 22 | 18 | 16 | 15 | 13 |
| | 336 | 332 | 257 | 189 | 148 | 114 | 89 | 72 | 58 | 53 | 49 |

**C.**

| mRNA expression | *NF1* high (n=4554) | *NF1* low (n=1112) |
|---|---|---|
| **Events**, n (%) | 2919 (64.1) | 673 (60.5) |
| **Median OS**, years | 3.56 | 4.19 |
| **5-Year OS rate**, % | 36.4 | 43.6 |

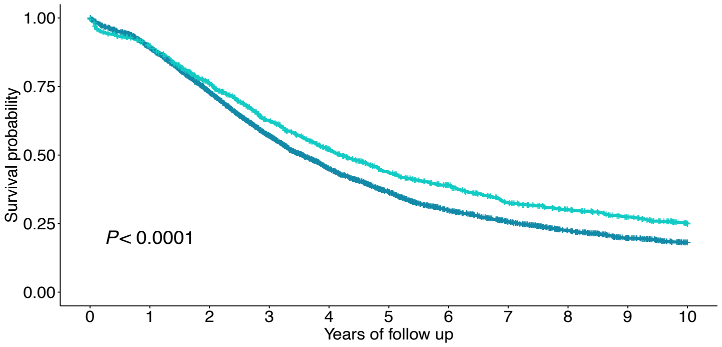


**Number at risk**

| | 0 | 1 | 2 | 3 | 4 | 5 | 6 | 7 | 8 | 9 | 10 |
|---|---|---|---|---|---|---|---|---|---|---|---|
| | 2587 | 3651 | 2970 | 2202 | 1598 | 1189 | 884 | 683 | 523 | 383 | 289 |
| | 520 | 892 | 766 | 620 | 498 | 391 | 322 | 253 | 212 | 171 | 123 |

**D.**

| | *BRCA1* & *NF1* high (n=133) | *BRCA1* & *NF1* low (n=44) | *BRCA2* & *NF1* high (n=70) | *BRCA2* & *NF1* low (n=36) | *BRCA*wt & *NF1* high (n=809) | *BRCA*wt & *NF1* low (n=216) |
|---|---|---|---|---|---|---|
| **Events**, n (%) | 64 (48.1) | 29 (49.6) | 42 (60) | 19 (52.8) | 573 (70.8) | 139 (64.4) |
| **Median OS**, years | 5.65 | 5.41 | 5.66 | 6.42 | 3.14 | 3.78 |
| **5-Year OS rate**, % | 54.3 | 56.5 | 54.3 | 70.3 | 32.6 | 39.9 |

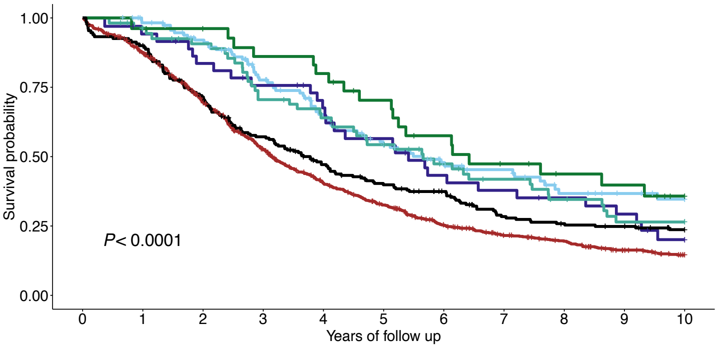


**Number at risk**

| | 0 | 1 | 2 | 3 | 4 | 5 | 6 | 7 | 8 | 9 | 10 |
|---|---|---|---|---|---|---|---|---|---|---|---|
| | 117 | 111 | 107 | 84 | 67 | 52 | 39 | 34 | 24 | 20 | 17 |
| | 33 | 35 | 32 | 30 | 25 | 22 | 16 | 14 | 12 | 10 | 6 |
| | 54 | 51 | 52 | 44 | 39 | 34 | 26 | 23 | 18 | 13 | 13 |
| | 26 | 26 | 28 | 28 | 26 | 22 | 17 | 14 | 11 | 10 | 6 |
| | 485 | 558 | 472 | 382 | 292 | 234 | 171 | 138 | 120 | 88 | 68 |
| | 125 | 139 | 131 | 118 | 97 | 82 | 72 | 55 | 51 | 48 | 39 |

### Extended Data Figure 8

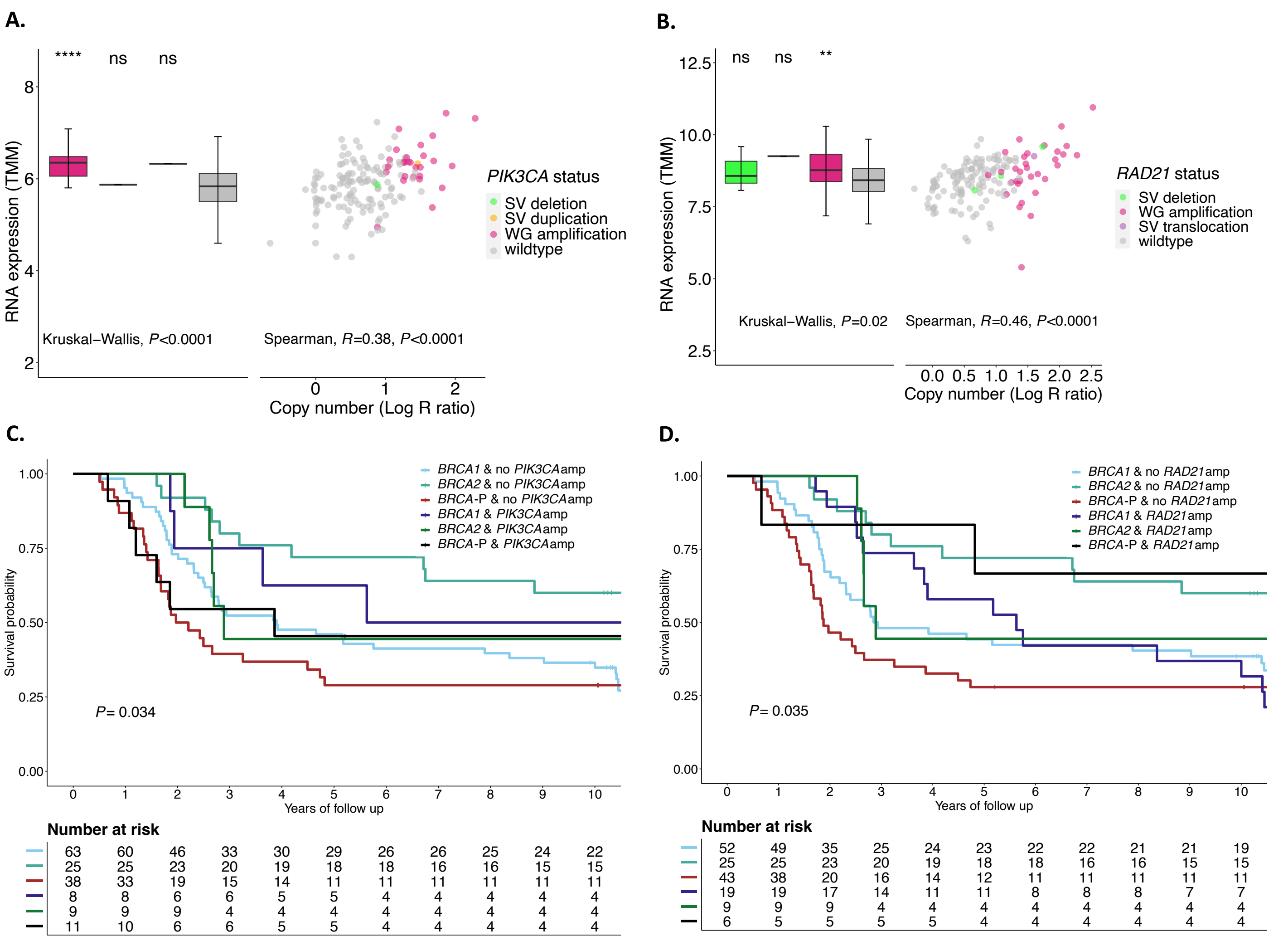


**E.**

| | BRCA1 & PIK3CA high (n=37) | BRCA1 & PIK3CA low (n=140) | BRCA2 & PIK3CA high (n=26) | BRCA2 & PIK3CA low (n=80) | BRCAwt & PIK3CA high (n=172) | BRCAwt & PIK3CA low (n=853) |
|---|---|---|---|---|---|---|
| **Events**, n (%) | 17 (45.9) | 76 (54.3) | 17 (65.4) | 44 (55) | 114 (66.3) | 598 (70.1) |
| **Median OS**, years | 7.67 | 5.23 | 4.09 | 7.43 | 3.21 | 3.22 |
| **5-Year OS rate**, % | 63.9 | 52.5 | 37.9 | 67.2 | 39.7 | 33.1 |

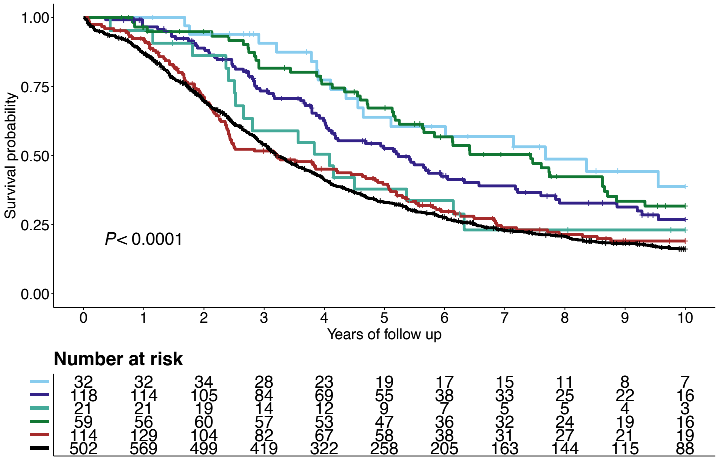

#### Extended Data Figure 9

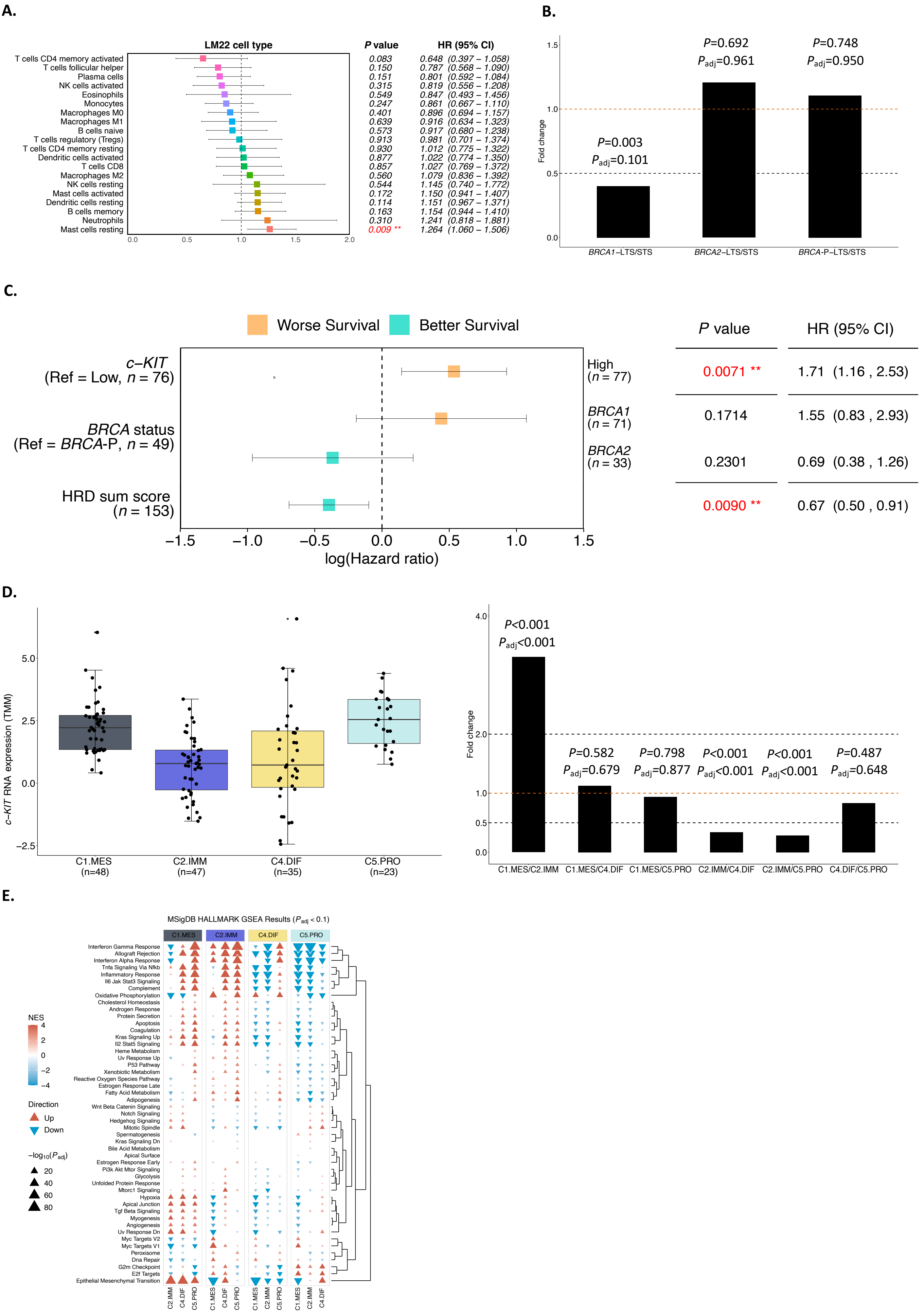
