## Supplementary Information for "Beyond *BRCA* deficiency: Clinical and molecular predictors of survival in patients with *BRCA*-deficient tubo-ovarian high-grade serous carcinoma"

1. Definition of survival groups
2. *BRCA*-deficiency
3. Sample quality control
4. Mutational signatures
5. NF1 immunohistochemistry analysis
6. Molecular subtype prediction
7. Clonality analysis
8. Mutual exclusivity
9. CIBERSORTx analysis
10. Methylation analysis
11. D11q analysis
12. Analysis of locus specific loss of heterozygosity in *NF1* alterations
13. Analysis of wildtype loss in *BRCA* mutated tumors

### 1. Definition of survival groups

The short overall survival (OS) cut-off of <3 years from diagnosis represented the lowest quartile of OS among germline pathogenic *BRCA* variant (g*BRCA*pv) carriers (n=281) in the AOCS cohort (**Supplementary Table S1, Extended Data Figure 1, Supplementary Information Figure 1A**). When restricted to only *BRCA* carriers who had died (n=170), three years remained a consistent cutoff to identify those in the lowest quartile of OS (**Supplementary Information Figure 1B**).

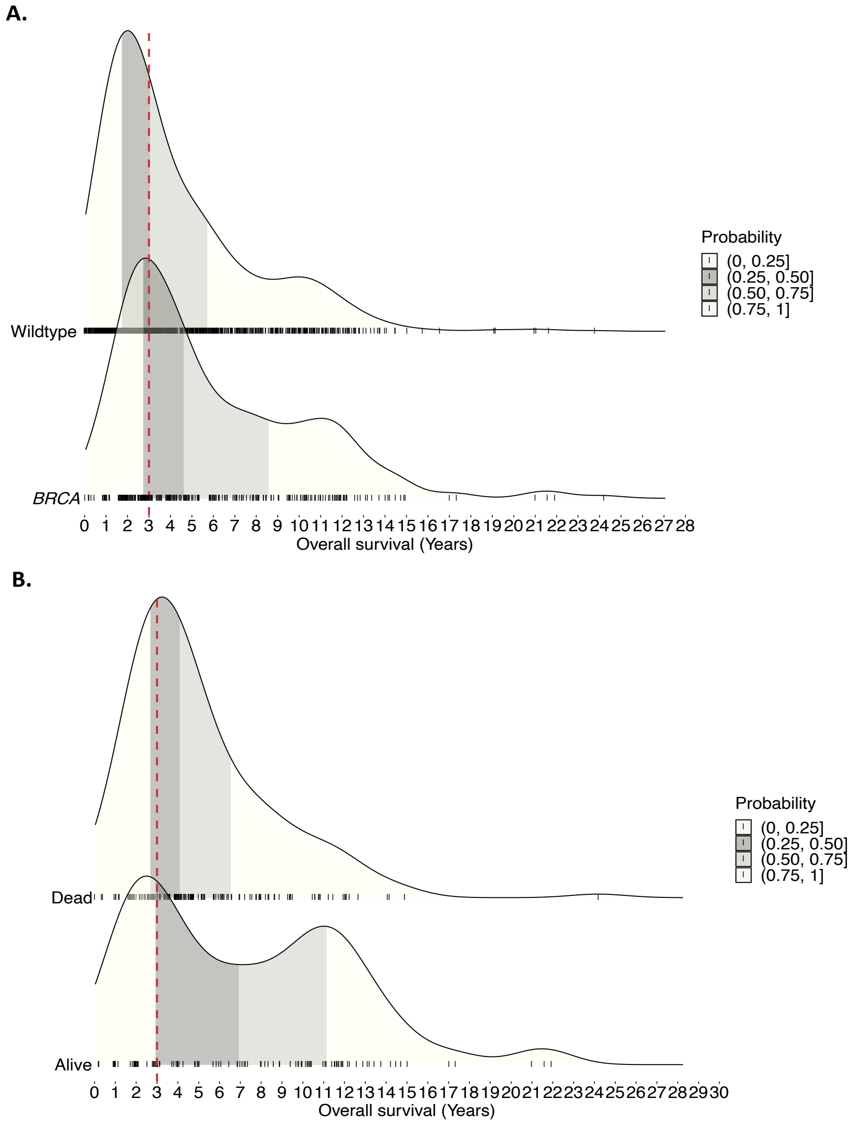


**Supplementary Information Figure 1 Survival distribution by *BRCA* status. A.** Density plots of overall survival for non carriers (n= 1096 *BRCA* wildtype; top) and patients with germline pathogenic *BRCA* variants (n=281 g*BRCA*pv; bottom), among patients with HGSC and known germline *BRCA* status in the AOCS cohort. **B.** Density plots of overall survival for deceased patients (n=170; top) and those alive at last follow-up (n=111; bottom), all of whom were diagnosed with HGSC and had g*BRCA*pv. The shaded areas represent different quartiles: 0.25-0.50 (ivory), 0.50-0.75 (gray), and 0.75-1.0 (light gray). The red dashed line marks the cut off of three years.

### 2. *BRCA*-deficiency

To classify tumors as *BRCA*-deficient, several of levels of evidence were considered: i) germline and tumor samples were evaluated for pathogenic or likely pathogenic alterations in the ovarian cancer risk and/or homologous recombination (HR) DNA repair genes *BRCA1* and *BRCA2* including promoter methylation of *BRCA1*, ii) pathogenic or likely pathogenic alterations in additional HR genes (*RAD51C*, *RAD51D*, *PALB2*, *FANCD2*, and *BRIP1*) were also evaluated, iii) the extent of genomic scarring associated with HR deficiency was assessed in tumor DNA using CHORD (CHORD score > 0.1). For analyses that required classifying samples as either *BRCA1* or *BRCA2* deficient, if there were multiple HR gene alterations in a tumor sample, the more dominant CHORD *BRCA1*- or *BRCA2*-type classification was selected. Tumor samples without an HR gene alteration and a CHORD score < 0.1 were classified as *BRCA*-proficient.

### 3. Sample quality control

Median tumor cell purity estimated by FACETS [1] was 0.63, with values ranging from 0.30 to 0.92 (**Supplementary Table S22**). BAMixChecker [2] was run on the paired Tumor/Normal WGS and RNAseq samples to look for sample concordance. One RNAseq sample "BRCA_5" was excluded because of its low concordance rate with the WGS data from the paired tumor sample.

### 4. Mutational signatures

We evaluated all mutational signatures from COSMIC (Catalogue Of Somatic Mutations In Cancer), SIGNAL, and Pan-cancer compendium. We then analyzed these mutational signatures in our cohort as previously described [3].

Signature fitting was conducted in two steps using the R package `signature.tools.lib` and its function `SignatureFit_withBootstrap`, following the methodology described previously [3]. Briefly, an initial fitting was performed using single-base substitutions (SBSs), double-base substitutions (DBSs), small insertions and deletions (IDs), copy number (CN) variations, ovarian cancer specific structural variants (SV) signatures, and chromosomal instability signatures which generated absolute signature exposures or contributions per sample, per signature. A second fitting focused on just ovary-specific signatures from COSMIC (for SBS, DBS, ID, and CN), SIGNAL (for SV), and the Pan-cancer compendium (chromosomal instabilities). Additional signatures present at high levels in the cohort were also included. Signatures were selected based on visualizing the mean signature exposures across the cohort and selecting an acceptable threshold. Absolute contributions were then converted into relative contributions by dividing each absolute value by the sample's total.

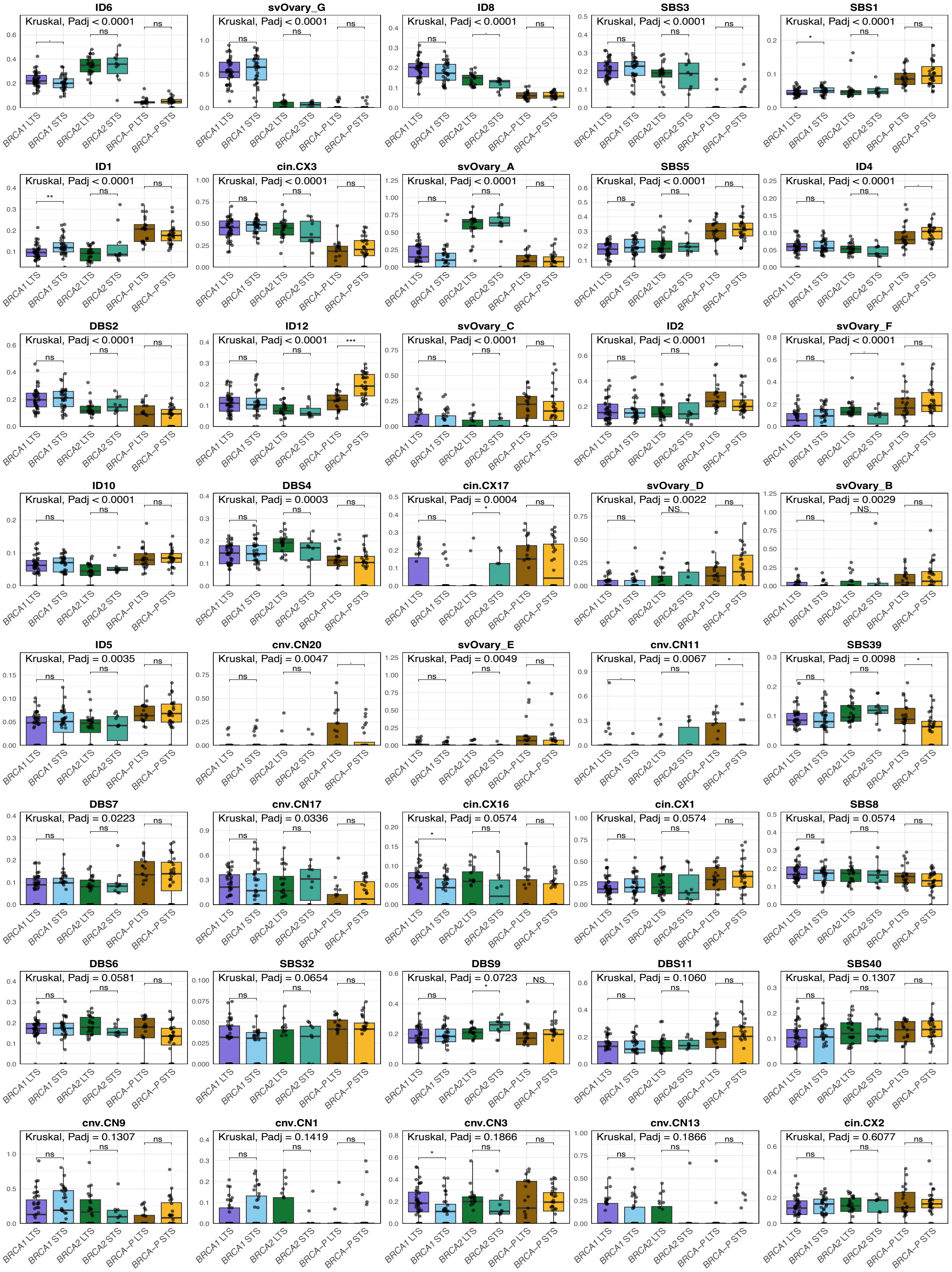


**Supplementary Information Figure 2 Distribution analysis of mutational signatures by *BRCA* and survival groups in HGSC. A.** Box plots summarizing mutational signatures by *BRCA* and survival groups; dots represent each sample, boxes show the interquartile range (25-75th percentiles), central lines show the median, whiskers show the smallest/largest values within 1.5 times the interquartile range. Kruskal-Wallis test

Benjamini-Hochberg adjusted $P$-values and pairwise Mann-Whitney-Wilcoxon test P values (two sided) are reported red (ns, $P>0.1$; ·, $P<0.1$; *, $P < 0.05$; **, $P < 0.01$; ***, $P < 0.001$). Features are ranked according to their significance.

### 5. NF1 immunohistochemistry (IHC) analysis

Tumor samples were analyzed for NF1 protein expression using IHC. The samples were classified into distinct categories: NF1 inactivated**,** where NF1 expression was absent in tumor cells but preserved in internal control tissues (e.g., stromal cells); NF1 retained, where normal NF1 protein expression was observed in the tumor cells; subclonal NF1 loss, indicating partial or patchy loss of NF1 expression in some but not all tumor regions; uninterpretable, where NF1 expression was absent in tumor cells but no internal control was available to validate the result; and excluded, where no viable tumor tissue was present for assessment (**Supplementary Information Figure 3**). Following this classification, survival analysis was conducted to compare OS in HGSC patients based on NF1 expression status (NF1 retained vs. NF1 loss). The analysis was also stratified by germline *BRCA* mutation status, allowing for a more refined evaluation of the interaction between NF1 status and *BRCA* mutations in determining patient outcomes. Results from this analysis are presented in **Extended data Figures 7A** and **7B**.

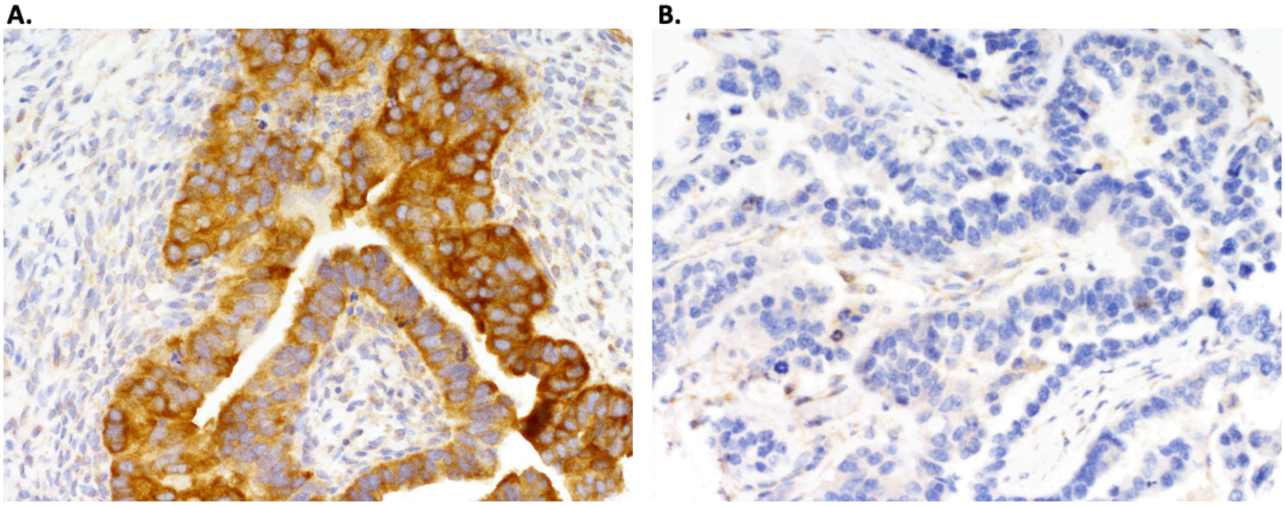


**Supplementary Information Figure 3** Representative immunohistochemistry images showing NF1 protein expression patterns in HGSC tumor samples. **A.** illustrates retained NF1 expression, with strong and uniform cytoplasmic staining observed in the tumor cells, indicating normal NF1 protein presence. The surrounding stromal tissue serves as an internal control with weaker staining. **B.** demonstrates complete loss of NF1 protein expression in the tumor cells, characterized by the absence of cytoplasmic staining, while stromal cells retain weak NF1 expression, serving as a reliable internal control for interpretation. These patterns were used to classify tumors as either NF1 retained or NF1 lost for further survival analysis.

### 6. Molecular subtype prediction

DeepCC (v0.1.1) [4] was used to classify the molecular subtypes of the tumor samples, using the Tothill dataset for training as previously described [3]. Briefly, to generate the training data, 1000 iterations were run to generate 1000 different models. This was done because each training run gave slightly different results when applied to the same test data. Molecular subtype predictions were then made by applying these models to the functional spectra of our batch-corrected RNA-seq data. The final molecular subtype was selected as the one that was predicted most frequently across the 1000 models.

### 7. Clonality analysis

Clonality estimation was performed as previously described [3]. Briefly, to obtain an estimate of the depth of the DNA repair pathway alterations and the number of variant reads, the alterations were manually checked in IGV. The total copy number at each variant locus was then determined by intersecting the alteration positions with the FACETS copy number data. The clonal or subclonal status of each variant was then classified by purity and copy number corrected cancer cell fraction (CCF). The CCF was calculated using the “absolute.cancer.cell.fraction” function [freely available at https://github.com/ucl-respiratory/preinvasive], by providing the total depth, variant read counts and FACETS total copy number for each alteration along with FACETS purity estimates for each sample.

### 8. Mutual exclusivity

Pairwise Fisher's Exact tests were conducted to assess the mutual exclusivity or co-occurrence of changes in genes of interest using the "somaticInteractions" function in maftools (v2.2.10) [3]. Only genes altered in at least three primary cancers were included in the analysis. The resulting *P*-values were adjusted for multiple hypothesis testing using the Benjamini-Hochberg method.

### 9. CIBERSORTx analysis

*Immune Cell Deconvolution:* We employed CIBERSORTx [5], a web-based tool available at https://cibersortx.stanford.edu/, for estimating immune cell populations within the tumor microenvironment. The analysis was configured with batch correction enabled, using absolute mode and 500 permutations, while quantile normalization was disabled. Deconvolution was based on the LM22 signature matrix.

*CIBERSORTx and Immunofluorescence Dataset Comparison:* The Spearman correlation test was employed to explore the relationship between CIBERSORTx results and immunofluorescence data. Specifically, we assessed the correlation between CD8+ T cell counts derived from the immunofluorescence data and the immune cell type specific absolute scores obtained from

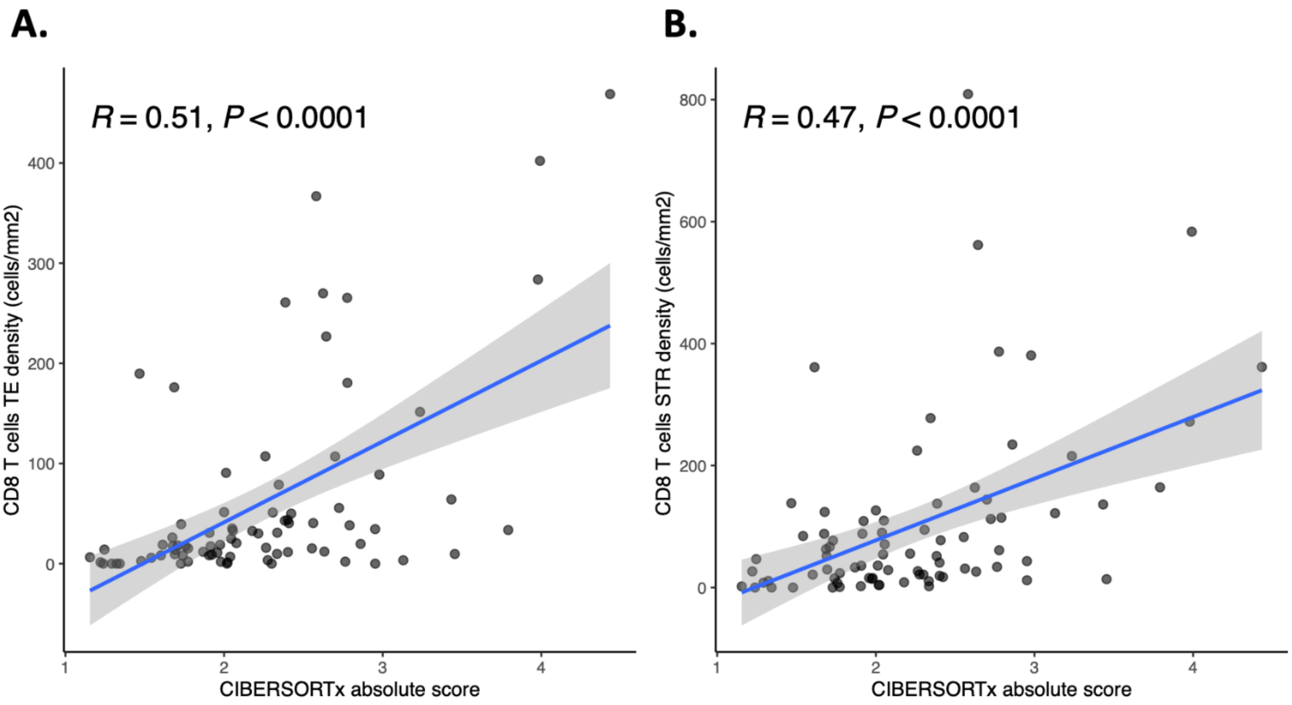


**Supplementary Information Figure 4 Comparison of RNA-seq-based immune cell estimation with immunofluorescence staining.** The correlation between CIBERSORTx absolute scores and CD8 T-cell densities was used to compare RNA-seq-based immune cell estimation with immunoflurescence staining. Scatter plots showing the relationship between CIBERSORTx absolute scores and CD8 T cell densities (cells/mm$^2$) for **A.** tumor epithelium (TE) and **B.** stroma (STR). In both plots, the blue line represents the linear regression fit and the shaded area indicates the 95% confidence interval. Spearman correlation *P* values are reported (two-tailed).

CIBERSORTx (**Supplementary Information Figure 4**). This analysis aimed to evaluate the consistency and reliability of CD8+ T cell quantification across both methods.

*Immune Cell Clustering:* Using the absolute cell abundance data from CIBERSORTx, we performed clustering using ConsensusClusterPlus [6].

1. Selection of Immune Cell Types: Only cell types with a nonzero value in at least 10 samples were included (**Supplementary Information Figure 5**).
2. Cluster Generation: After selecting the cell types as per the criteria above, the sample by cell type matrix of absolute enrichment scores we used to generate the immune cell clusters following a methodology similar to the mutational signature cluster analysis.

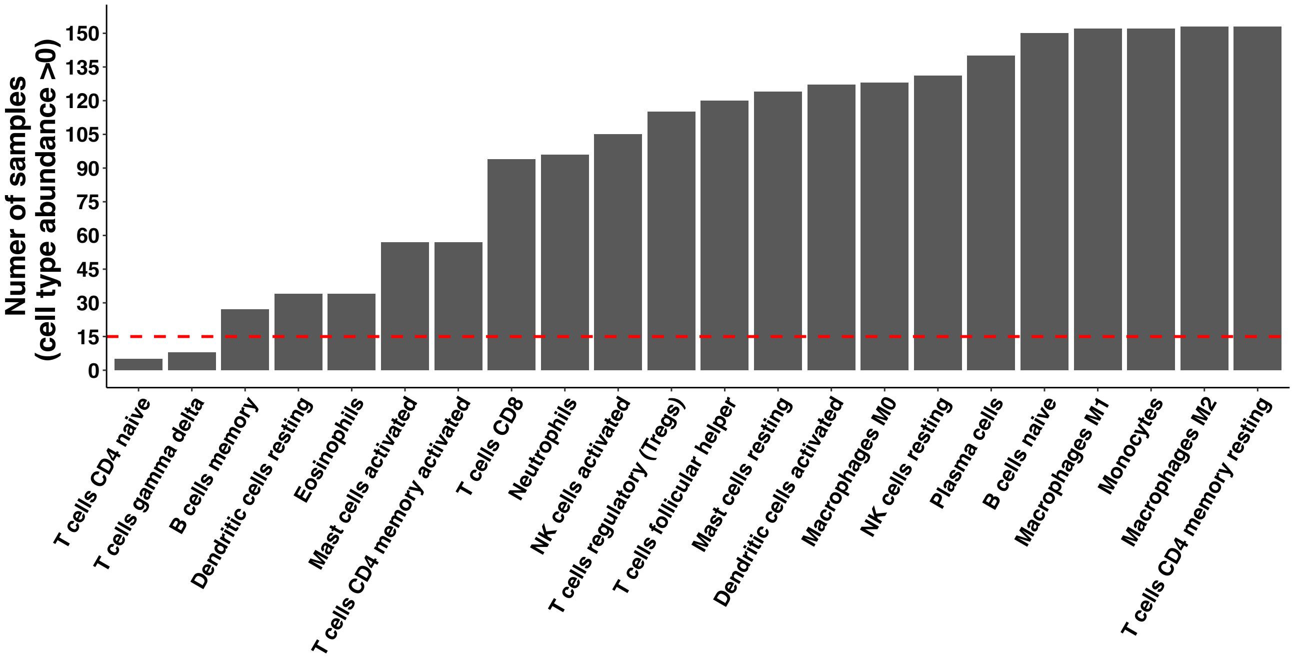


**Supplementary Information Figure 5 Cell type selection for use in clustering analysis.** Bar chart showing the distribution of cell type abundances from CIBERSORTx analysis. The x-axis lists all immune cell types. The y-axis represents the number of samples in which each cell type is detected with an abundance greater than zero. This visualization highlights the prevalence of different immune cell types across the analyzed samples. The cut-off for cell types included in the clustering is indicated by the red dashed line.

### 10. Methylation analyses

*Methylation classification of gene promoters:* To determine the methylation status of the *BRCA1* and *RAD51C* gene promoters, we analyzed their mRNA expression levels in relation to the methylation array beta values of probes situated in the 5’ CpG island of each gene. This analysis was conducted across all primary tumor samples (n = 153) using previously established methods [3]. To determine the methylation status of the *BRCA1* and *RAD51C* gene promoters, we analyzed their mRNA expression levels with respect to to the methylation array beta values of probes located in the 5' CpG island of each gene. This analysis was performed on all primary tumor samples (n = 154) using previously established methods. Specifically, we selected methylation probes that showed a significant negative correlation with mRNA expression (Spearman's r < -0.30, P < 0.01). For *BRCA1*, 15 probes (cg04110421, cg04658354, cg08386886, cg09441966, cg09831010, cg10893007, cg15419295, cg16630982, cg16963062, cg18372208cg, 19088651, cg19531713, cg20187250, cg21253966, and cg24806953), and *RAD51C*, 2 probes (cg02118635 and cg24099023) were selected. Samples were classified as methylated if their beta values exceeded 0.2 in at least 50% of the selected probes and/or the samples were classified as methylated as previously analyzed with the same method on the 126 primary tumors included in the previous study [3].

*Differential Methylation vs Differential Expression analysis*

Differential Methylation (DM): A combined beta matrix of 154 patients and 364,185 probes was generated by intersecting the common probes between the 450k and EPIC methylation assays. The beta values were converted to M-values [7] and the R package Limma (v3.48.2) was used to identify differentially methylated probes between the groups of interest. A batch effect was observed using multidimensional scaling (MDS) plots between the methylation assays and was subsequently included in the model during the DM analysis.

Differential Expression (DE): Please refer to section "*Differential expression analysis*" in the main methods.

We used starburst plots to assess the interaction between methylation and expression [8].

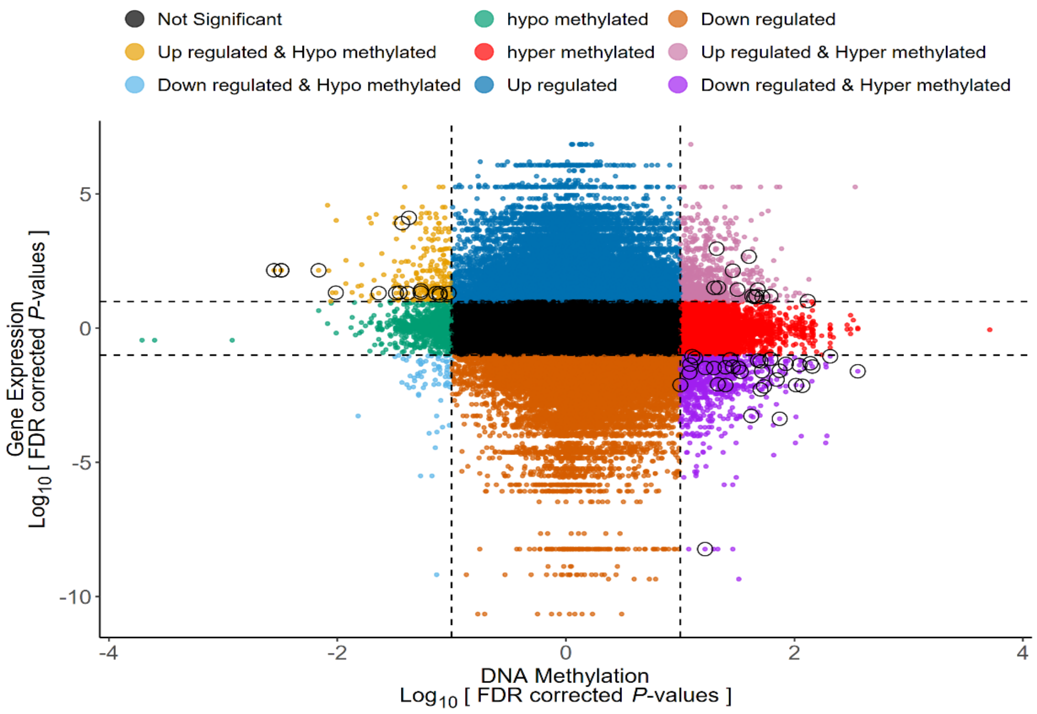

To generate data for the starburst plots, the $P_{adj}$ values from the DM and DE analyses were converted to $-\log_{10}(P_{adj})$ values and the direction of the fold changed was applied to these values (signed $P_{adj}$ values). The signed $P_{adj}$ values were then plotted along the X-axis for methylation, and Y-axis for expression. The dotted lines indicate a $P_{adj}$ value of < 0.1 and the colored dots indicate various groups of interaction between the methylation and expression data. The black dots in the center denote genes and probes with no significant interaction in the DM vs DE analysis (annotated "Not Significant" in the legend). The circled dots are probes that are significant in both the DM and DE analysis with an ADB (Absolute Delta Beta) of > 0.2 to indicate a notable difference in the mean beta values between the contrasting groups. Only significant probes with $P_{adj} < 0.1$ for both DM and DE analyses that fell in the top-left (Up regulated & Hypo methylated), top-right (Up regulated & Hyper methylated), bottom left (Down regulated & Hypo methylated) and bottom-right (Down regulated & Hyper methylated) were output to the results tables (**Supplementary data**). We then focused on the bottom right and top left portion of the starburst plot i.e. the group of genes that were significantly ($P_{adj} < 0.1$) "Down regulated & Hyper methylated" or "Upregulated & Hypo methylated" with ADB > 0.2 (**Supplementary Table S17**).

**11. D11q analysis**

*Creation of d11q Expression Data:* Reads mapping to BRCA1 coordinates (GRCh38 17:41196312-41277468) were extracted into individul BAM files for each sample (n=153). The BAM files and a GTF containing the canonical and d11q BRCA1 gene models were then fed into the R library ASpli (v2.0.0) to generate splice junction counts. The d11q splice junction proportions were calculated as [d11q_splice_junction_counts/(d11q_splice_junction_counts + canonical_splice_junction_counts)] where "d11q_splice_junction_counts" were reads mapping to the d11q splice junction and "canonical_splice_junction_counts" were reads mapping to the canonical splice junction.

*Selection and Analysis of d11q Expression:* The d11q expression data was filtered using a threshold of 10 for d11q exon counts. After applying this threshold, 122 samples remained for the final analysis (**Supplementary Table S12)**. The selected samples were then subjected to further statistical and bioinformatic analyses to explore the expression patterns and their potential implications in the study (**Figure 2B-E)**.

**12. Analysis of the locus specific loss of heterozygosity in *NF1* alterations**

To assess the locus specific loss of heterozygosity (LOH) in *NF1* alterations, we integrated tumor sequencing data, including variant and reference allele read counts, variant allele frequencies (VAFs), clonality, and lower copy number (LCN) (**Supplementary Table S18**). Tumor VAFs were calculated as the proportion of variant allele reads to total reads. These observed tumor VAFs were then compared to the expected values based on LCN and clonality. Locus specific LOH was indicated in tumors with reduced copy number (LCN = 0). Discrepancies between observed and expected VAFs were evaluated for potential contributions from tumor purity, technical artifacts, or additional copy number alterations. Locus specific LOH was determined by concordant evidence from elevated tumor VAFs, copy number status, and clonality. This classification was further supported by the depletion of mRNA expression in tumors with NF1 alterations exhibiting locus specific LOH ($P<0.0001$; **Extended Data Figure 6A**).

**13. Analysis of wildtype loss in *BRCA* mutated tumors**

To assess the loss of the wildtype allele in *BRCA* cases, we integrated germline and tumor sequencing data, including variant and reference allele read counts, VAFs, clonality, and LCN (**Supplementary Table S15**). Germline and tumor VAFs were calculated as the proportion of variant allele reads to total reads. Germline VAFs were validated to confirm heterozygosity, typically around 50%.

The tumor VAFs were compared to the expected values based on LCN and clonality. For tumors with normal copy number (LCN = 1), retention of the wild-type allele was indicated by a tumor VAF approximating the germline value, while loss of the wild-type allele (LOH) was inferred from VAFs approaching 100%. For reduced copy number (LCN = 0), tumor VAF near 100% indicated loss of the wild-type allele.

While LCN was used as the primary metric for assessing allelic loss, total copy number (TCN) data provided additional context, with TCN = 1 suggesting loss of a single allele and TCN = 0 indicating homozygous deletion. Discrepancies between observed and expected VAFs were evaluated for potential contributions from tumor purity, technical artifacts, or additional copy number alterations. Loss of the wild-type allele was determined by concordant evidence from elevated tumor VAFs, copy number status, and clonality.
